# Modeling COVID-19 disease processes by remote elicitation of causal Bayesian networks from medical experts

**DOI:** 10.1101/2022.02.14.22270925

**Authors:** Steven Mascaro, Yue Wu, Owen Woodberry, Erik P. Nyberg, Ross Pearson, Jessica Ramsay, Ariel Mace, David Foley, Tom Snelling, Ann E. Nicholson, COVID BN Advisory Group

## Abstract

**Background:** COVID-19 is a new multi-organ disease, caused by the SARS-CoV-2 virus, resulting in considerable worldwide morbidity and mortality. While many recognized pathophysiological mechanisms are involved, their exact causal relationships remain opaque. A better understanding is needed for predicting their progression, targeting therapeutic approaches, and improving patient outcomes. While many mathematical causal models describe COVID-19 epidemiology, none have been developed for its pathophysiology. The virus’s rapid and extensive spread and therapeutic responses made this particularly difficult. Initially, no large patient datasets were publicly available, and their data remains limited. The medical literature was flooded with unfiltered, technical and sometimes conflicting pre-review reports. Clinicians in many countries had little time for academic consultations, and in-person meetings were unsafe.

**Methods and Findings:** In early 2020, we began a major project to develop causal models of the pathophysiological processes underlying the disease’s clinical manifestations. We used Bayesian network (BN) models, because they provide both powerful tools for calculation and clear maps of probabilistic causal influence between semantically meaningful variables, as directed acyclic graphs (DAGs). Hence, they can incorporate expert opinion and numerical data, and produce explainable results. Dynamic causal BNs, which represent successive “time slices” of the system, can capture feedback loops and long-term disease progression.

To obtain the likely causal structures, we used extensive elicitation of expert opinion in structured online sessions. Centered in Australia, with its exceptionally low COVID-19 burden, we managed to obtain many consultation hours. Groups of clinical and other subject matter specialists, all independent volunteers, were enlisted to filter, interpret and discuss the literature and develop a current consensus. We aimed to capture the experts’ understanding, so we encouraged discussion and inclusion of theoretically salient latent (i.e., unobservable) variables, documented supporting literature while noting controversies, and allowed experts to propose mechanisms by extrapolation from other diseases. Intermediary experts with some combined expertise facilitated the exchange of knowledge to BN modelers and vice versa. Our method was iterative and incremental: we systematically refined and checked the group output with one-on-one follow-up meetings with the original and new experts to validate previous results. In total, 35 experts contributed 126 face-to-face hours, and could review our products.

**Conclusions:** Our method demonstrates and describes an improved procedure for developing BNs via expert elicitation, which can be implemented rapidly by other teams modeling emergent complex phenomena. The results presented are two key models, for the initial infection of the respiratory tract and the possible progression to complications, as causal DAGs and BNs with corresponding verbal descriptions, dictionaries and sources. These are the first published causal models of COVID-19 pathophysiology, with three anticipated applications: (i) making expert knowledge freely available in a readily understandable and updatable form; (ii) guiding design and analysis of observational and clinical studies, by identifying potential mediators, confounders, and modifiers of treatment effects; (iii) developing and validating parameterized automated tools for causal reasoning and decision support, in clinical and policy settings. We are currently developing such tools for the initial diagnosis, resource management, and prognosis of COVID-19, parameterized using the ISARIC and LEOSS databases.

## 1 Introduction

COVID-19 is a new multi-organ disease, caused by the highly infectious SARS-CoV-2 virus, resulting in considerable worldwide morbidity and mortality. While many recognized pathophysiological mechanisms are involved—coagulation and inflammatory cascades, pulmonary exudation and respiratory compensation, endovascular and renal injury—the exact causal relationships between them remain opaque. A better understanding of these causal processes is urgent and important for predicting their progression, targeting therapeutic approaches and improving patient outcomes. To this end, the worldwide pandemic generated a flood of research, but much of the early output was unfiltered, sometimes conflicting, and of reduced reliability [1]. In contrast, no large patient datasets were publicly available, and even now, their data remains limited.

In this project, we used online tools to enlist independent medical experts to interpret and iteratively discuss the evolving literature, providing us with consensus views. This process resulted in the elicitation of several detailed graphical causal models of the pathophysiology underlying the clinical manifestations of COVID-19 disease, using the kind of iterative, incremental method widely recommended for model building [2], but rarely realized in expert knowledge elicitation. From these theoretical products, simplified models can be constructed for practical application that are parameterized using available data, e.g., for predicting the probability of a patient’s future need for intensive care given their current signs, symptoms and laboratory results.

In this paper, we make freely available our expert-elicited models. While there has been considerable prior work developing causal models of the transmission of SARS-CoV-2 within populations [3–11], ours are the first causal models of the COVID-19 disease process within individuals. We anticipate that these models will serve three purposes, for our team and other researchers. First, they will aid communication and theoretical understanding, which includes being readily updated to incorporate new findings, and being internally and externally validated using local datasets. Second, they will guide the design and analysis of both observational and clinical studies into COVID-19, by identifying potential mediators, confounders, etc. Third, they will be used to develop and validate parameterized practical tools for causal reasoning and decision support, both in clinical and policy settings.

### 1.1 Causal Bayesian networks

Bayesian networks (BNs) were designed to model probabilistic causal systems. Formally, a BN [2, 12, 13] includes a directed, acyclic graph (DAG), (e.g., Fig. 1), which consists of nodes connected by arcs (pointing from ‘parent’ to ‘child’) that never point in a continuous sequence from a node back to itself.^1^ In addition, in a BN each node represents a random variable with multiple possible states, and each arc represents a direct probabilistic dependency, quantified by a conditional probability table (CPT) or equation specifying the probability distribution of the child given the states of all its parents. In *causal* DAGs and BNs, these arcs also represent direct causal influences—hence they can also predict the effects of decisions regarding interventions.

**Fig 1.**
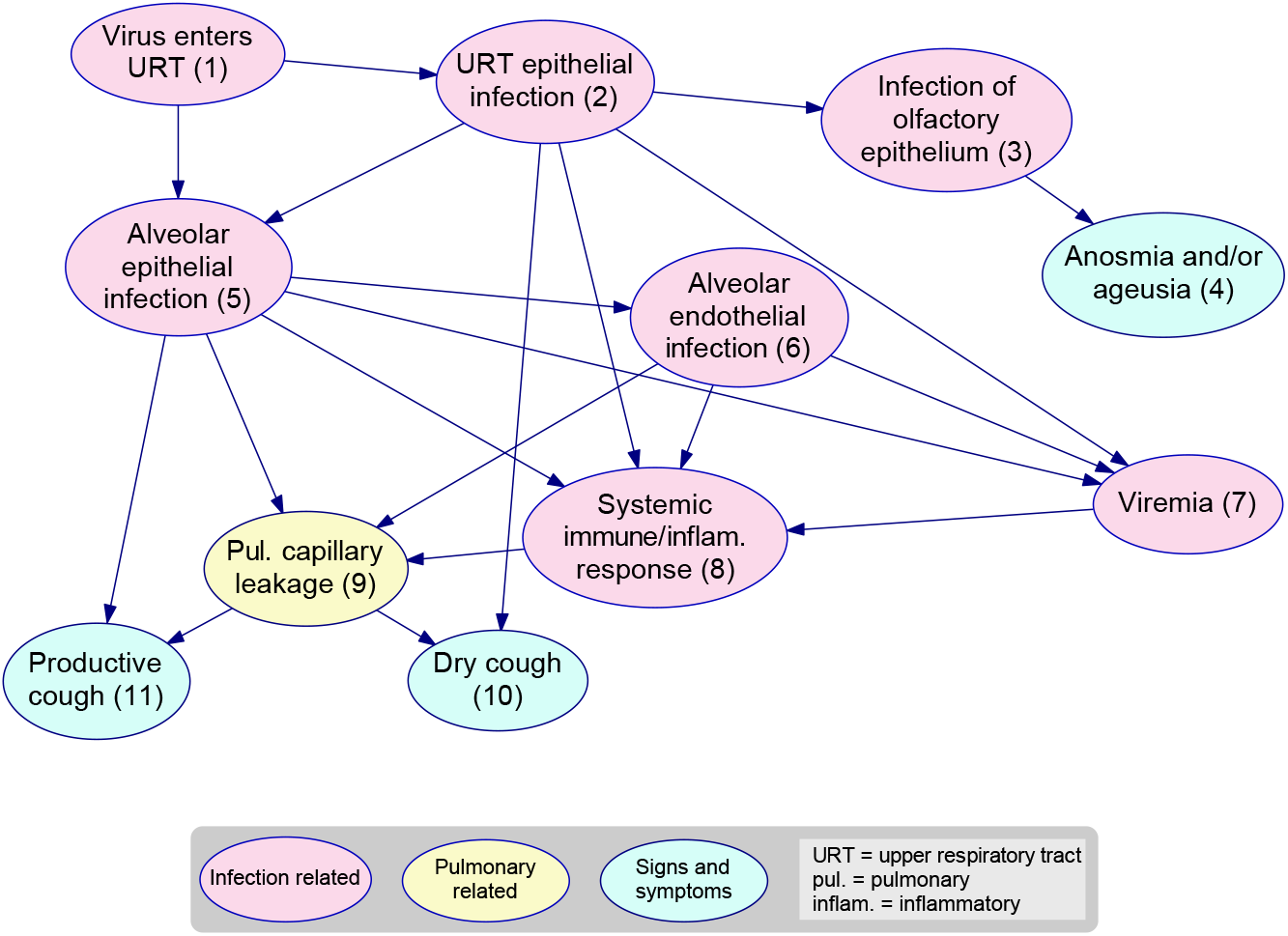
Causal DAG excerpt. These are the initial nodes of the *Respiratory* causal DAG provided in S1 Fig. The node Pulmonary capillary leakage, for example, has three ‘parents’ that jointly influence it, and two ‘children’ it influences. In the BN, provided in S1 Model, the variable it represents has states and a CPT that are not shown in the DAG. All the nodes are named, numbered, and color coded for ease of reference, and corresponding definitions for variables and arcs are given in the associated Dictionary, S1 Table, an excerpt from which is provided in Fig. 2.

Readily available software^2^ allows users to enter exact or uncertain evidence about any variables, which is then efficiently propagated to update the probability distributions for all variables. Thus, causal BNs can support and perform prognostic (predictive), diagnostic (retrodictive), explanatory and decision-oriented probabilistic reasoning.

In a BN, all arcs and nodes—even ‘latent’ ones, i.e., not directly observable—may be defined in a semantically meaningful way (as in Fig. 1 and Fig. 2), and this will always be so when the BNs represent knowledge elicited from humans. This transparency is in stark contrast to the opaque “black boxes” of neural networks, where latent nodes are usually connected and parameterized based on a large dataset and do not have any readily discernible semantic or causal interpretation.

**Fig 2.**
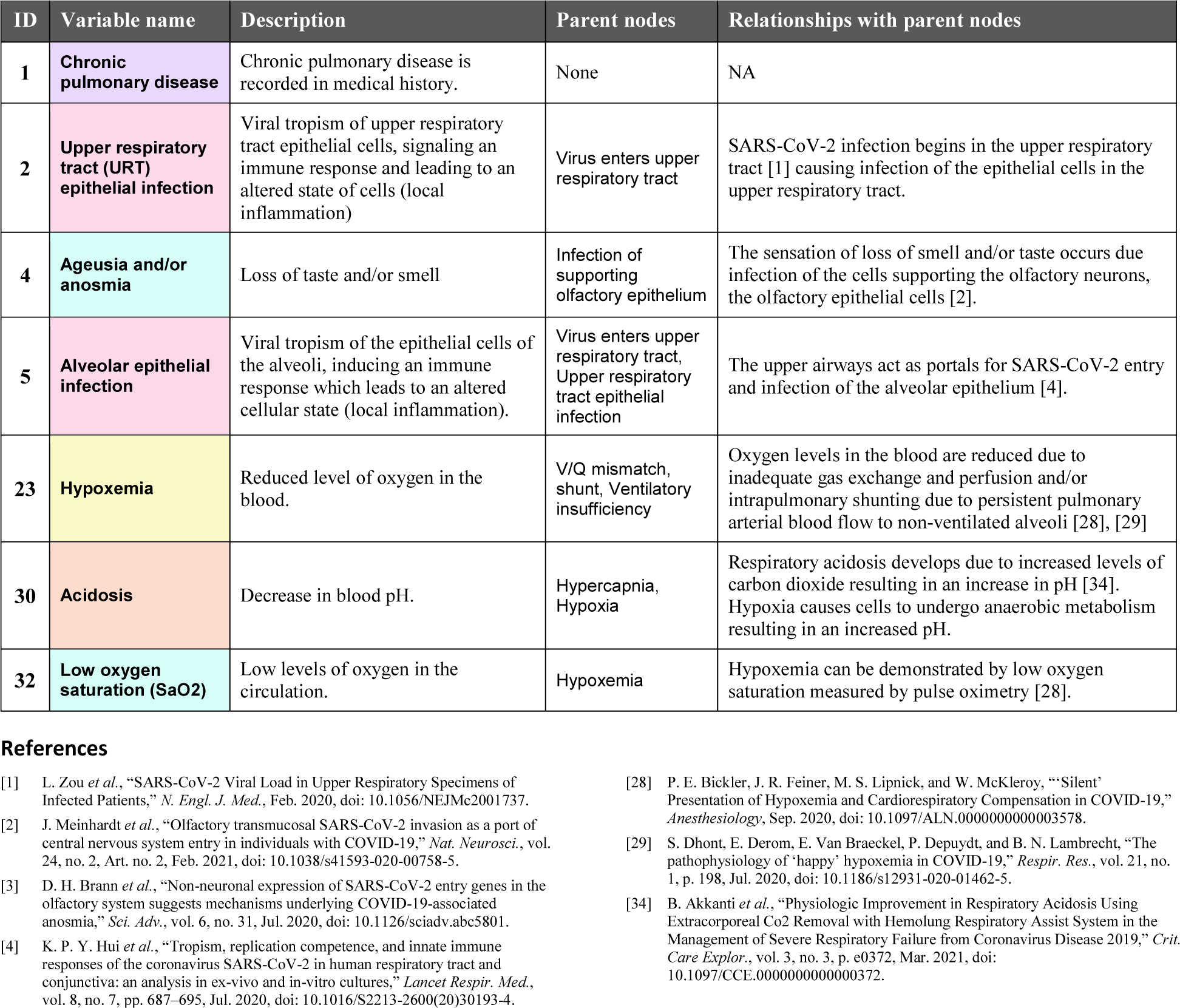
BN Dictionary excerpt: In the Dictionaries, descriptions of the variables and arcs are provided, with selected supporting references. The variables are numbered, named and color coded as in the causal DAGs and BN files. The full Dictionaries are provided as S1 Table and S2 Table.

One important consequence is that an elicited BN can be readily updated to reflect new human knowledge, which is often derived from more than just a new dataset. Since it is clear which nodes the new knowledge relates to, we can usually identify a few locations where changes are needed while leaving the remainder of the network untouched. This can be extremely useful for complex emergent phenomena, where understanding is advancing rapidly while datasets remain small.

Another important consequence is that the BN structure and its inferences are potentially explainable, provided that the important aspects of a complicated BN or inference can be identified and clearly articulated. In medicine, for example, for ethical and practical reasons it is better if the recommendations of an AI tool are explainable and justifiable [14, 15]. Algorithms for automatically generating such explanations, which supplement basic BN software by distilling complex inferences into a concentrated essence more palatable for human consumption, are a continuing focus of research, e.g., [15–19]. These explanations are often better expressed in causal terms [20], including those supporting medical diagnoses [21].

Any causal BN entails a partial time order for its variables: parents precede their children. But a BN’s capacity to represent time is limited, since cycles are prohibited and each variable usually appears only once, thus making it difficult to represent both feedback loops and time series. A dynamic Bayesian network (DBN) [2] is a BN in which a variable can be represented multiple times at different discrete time steps, and at any step *T* it may have parents at *T, T* − 1, *T* − 2, *T* − 3, … Hence, DBNs can model any complex multivariate time series, including feedback loops. If the structure of the network remains constant over time, the DBN can be compactly and clearly represented as a network “snapshot” at a time step showing each variable’s connections to all its parents.

We present our models as non-dynamic BNs by focusing on the first instance at which each variable becomes involved, but have noted some feedback loops to facilitate their possible conversion to DBNs for modeling disease progression over longer time scales.

### 1.2 Building BNs by eliciting expert opinions

Given a large and representative numerical dataset, preferably supplemented by expert guidance on some causal directions, machine learning algorithms have been developed to find the sparsest causal BNs that could produce the observed dependencies, e.g., PC [13], CaMML [22], and R libraries such as bnlearn (https://www.bnlearn.com/). In the absence of such a dataset, BN structure and/or parameters—unlike those of opaque models—can be obtained entirely through knowledge elicitation from domain experts. This may be more time consuming, but has the essential advantage that expert knowledge usually represents a synthesis of multiple sources that can supplement and help interpret limited local data. Since domain experts usually have no prior exposure to BNs or training in causal inference, they need to be assisted by an expert BN modeler, who helps to express the knowledge in appropriate terms and highlight any important or missing pieces.

It is common to use multiple opinions rather than relying on only one source, with the hope of obtaining a larger pool of information and greater reliability from aggregated opinions, and perhaps benefiting from mutual feedback via discussion. Group elicitation of BNs has been particularly popular in some domains, such as reliability assessment [23, 24] and environmental research [25].

When group knowledge is pooled effectively, whether by collation, consensus, averaging, or another form of amalgamation, then it tends to outperform individual knowledge [26–31]. However, unstructured group interactions also bring well-documented problems, e.g., diffusion of responsibility, anchoring, ‘groupthink’, deference to social status, and unproductive disputes [32–36]. Such problems can be reduced by using structured protocols, e.g., collecting independent opinions from all participants prior to discussion, and also by using a moderator, e.g., to focus discussion on the most pressing issues, encourage only constructive contributions, and help establish a consensus where possible. A BN modeler with appropriate skills may also take the role of the moderator who leads discussion, which is the approach we used.

Software tools have recently been developed to support group elicitation of BNs, and the BARD tool [19] includes multiple options to support highly structured protocols, including a strict version of the widely used Delphi technique [37, 38]. However, we needed to make the elicitation interactions as easy and quick as possible for our experts, so using only a videoconferencing tool (Zoom) with a modeler/facilitator who followed structured but flexible protocols was the most appropriate choice.

The way BNs are defined in § 1.1 might suggest that we build them in the same logical stages, like a waterfall: (i) define all the variables, so that we can (ii) specify all the arcs between them, and then (iii) estimate all the parameters this structure requires. However, for complex BNs this would require uncanny foresight. In contrast, as noted in [19], proposed methodologies for BN elicitation recommend proceeding iteratively and incrementally [2, 39–41]. More specifically, [2, Part III] suggests beginning with a small local structure around a target variable of interest, rather than attempting to exhaustively consider every possible factor relevant to the target. Subsequent iterations can pick up a few additional factors at a time, preferably with some form of validation in each iteration (e.g., feedback from an independent expert).

Another recommended strategy is to break down complex models into sub-models, and reuse common structures or elements when appropriate, dubbed “idioms” [42], “templates” [40] and “network fragments” [39]. These incremental approaches adapt similar ideas long used in software engineering, such as “spiral prototyping” or “agile model building” [43], and reusing common local structures is fundamental to “object-oriented” programming [44]. An expert modeler/moderator can manage this workflow flexibly and efficiently as issues emerge.

If probabilities are elicited, then there is usually some degree of uncertainty and disagreement about them. Instead of a single point estimate for each probability, more thorough protocols have been designed to elicit several points (e.g., maximum, minimum, and best estimates) from each of multiple experts and combine this information [45–48]. Here, we are concerned principally with causal structure, with probabilities to be determined later from numerical datasets—although frequently experts provide rough indications of influence along with the variables and causal arcs, which can be captured by ‘indicative’ BN parameters and used for structure validation (see § 2.3).

### 1.3 Our contributions

This paper makes two kinds of novel contributions to prior literature: in method and in results. In § 2, we demonstrate a structured approach that is only feasible using online tools, which manages to combine large-scale group expert elicitation with BN building that is iterative, incremental, and includes validation. In § 3, we present detailed causal DAGs and BNs (with indicative parameters), together with comprehensive associated documentation, that capture our experts’ theoretical understanding of the corresponding COVID-19 pathophysiological processes. In § 4, we explain how researchers can use both our methods and results, and how we are currently doing so.

## 2 Methods

Our elicitation process was organized very quickly, relative to its scale and to similar projects with which any of the present authors have been involved. Organization included our initial planning for which models we wanted to develop, and hence what information we needed to elicit.

The process was exceptionally intensive, with many experts involved, a high degree of interest, and more detail captured in the expert models than usual. Centered in Australia, with its exceptionally low COVID-19 burden, we managed to obtain a very large number of consultation hours—mostly from experts around Australia, but supplemented by experts overseas. Many of these clinical experts therefore had little direct experience in treating COVID-19, with the exception of some overseas recruits with firsthand experience. Nevertheless, since their role was to interpret the available literature in the context of their existing domain knowledge rather than contribute original material, their expertise was adequate for our purposes. Furthermore, in high incidence settings few experts would have had the time to participate.

There was a high degree of consensus about what was currently established, except when discussing immune processes. However, knowledge in this area was evolving rapidly as new evidence emerged, so as expert opinion was revised we needed to update our models.

### 2.1 Social roles for elicitation

There are three major specialist roles people performed during the elicitation process, which we will refer to as follows:

**Modelers** are our team’s technical experts in BN and other computational modeling and causal inference, which encompasses model construction, refinement, comparison, learning via expert elicitation, machine learning from data, validation, and application.

**Medical Experts** are the domain experts we enlisted to contribute their time and knowledge, which here means medical specialists in a relevant subsystem, who agreed to participate in any of our elicitation activities.

**Coordinators** are people on our team with some combination of general medical knowledge, general modeling knowledge, and medical contacts, who facilitated interaction between modelers and experts, and searched the available literature to support the model-building process.

Since both medicine and modeling are highly technical, our cross-disciplinary coordinators played a vital role in translating knowledge from one sphere to the other. Also, they ensured that the modeling decisions our team made were always informed by general medical expertise even where we did not elicit specialist medical opinions on the issue.

Some members of our team (YW, TS and SM) had previous experience in building causal models for infectious disease pathophysiology, including both a generalizable basic structure and adaptations for specific cases such as pneumonia. Based on a preliminary literature review, we adapted this generalizable model to form a preliminary model for COVID-19.

However, due to the recent emergence of the disease, compounded by its inherent novelty and complexity, it quickly became clear that obtaining the necessary causal knowledge would be a massive challenge. First, there was insufficient data available in the published literature to infer causal mechanisms, and the available literature was highly specialized and fragmented. Second, there was insufficient raw data available to infer causal structure more directly using machine learning techniques (such as those mentioned in § 1.2). Third, there was insufficient certainty amongst experts to rely on any individual source of authority; it would be more reliable to build an understanding from a group consensus, such as a group of experts in respiratory medicine. Fourth, the disease involves multiple areas of specialist knowledge, so we would need to elicit opinions from multiple groups of medical experts. Fifth, the logistical challenge of enlisting and managing multiple groups would only be compounded by needing to conduct all meetings online, due to the social effects of the very disease under discussion. Lastly, several models of specific subsystems would need to be synthesized into a single, general master model to provide the theoretical foundation for developing clinical tools. Nevertheless, we took on this difficult challenge.

### 2.2 Initial models

How to divide the problem into manageable models and associated groups of experts was not obvious, since medical expertise is differentiated by more than one dimension. Some medical specialists focus on particular body systems, and here early clinical experience indicated that COVID-19 strongly affected the respiratory and immune systems, with frequent complications, often involving the hematological, cardiovascular and renal systems. Clinicians, however, are familiar with the processes of diagnosis and prognosis, which need to synthesize the most important information from all these physiological systems. Informed by our previous modeling experience and preliminary literature review, we initially divided the problem into four submodels: *Core Mechanism, Complications, Immune Response*, and *Diagnosis*. (This division was eventually revised substantially, as described in § 3.1.) Our *Core Mechanism* submodel was focused primarily on the respiratory system, so we primarily enlisted respiratory and infectious disease specialists to help create it. *Immune Response* details were elicited from clinical immunologists and other domain experts in human immunology. In contrast, to discuss *Complications*, we enlisted a more varied group of clinicians and other domain experts, including cardiologists, intensivists, and renal specialists. Our experts for *Diagnosis* were also more varied, including emergency and primary care physicians, and medical microbiologists.

To help orient our medical specialists, we provided a simple schematic *Overview* model indicating how these four submodels were related. However, in addition to developing the four submodels, we needed to actually synthesize them into a unified model with a view to developing practical tools to facilitate clinical reasoning and decision-making. This task was performed by our modelers in parallel, i.e., as the submodels were refined, so was the unified model.

We were able to leverage the insights from submodel workshops to connect the submodels appropriately, and to prioritize the most important features and omit less important ones. We did not use separate elicitation sessions for developing the unified model, because it was constrained to be consistent with the four submodels, it was too complex to address in a single workshop, and it also involved decisions based on modeling expertise.

### 2.3 Elicitation sequences

Fig. 3 illustrates our use of several types of elicitation session in a specific sequence, and how these sessions mapped to the sequence of steps needed to develop a model.

**Fig 3.**
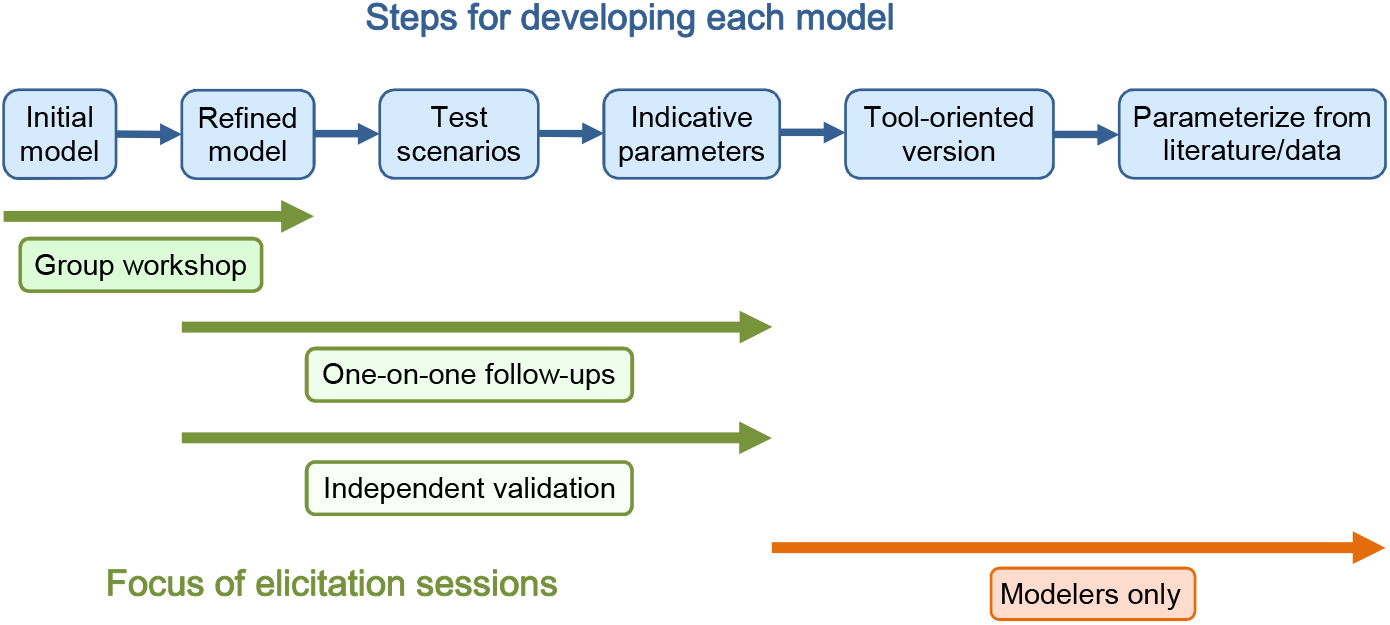
Typical steps used in our model development process, and their relationship to our expert elicitation sessions.

Our first elicitation session to develop a particular expert causal model was a 2 h group workshop, with 7**–**12 experts each attending for 1**–**2 h. The main goal here was to develop a causal structure and refine it through interactive discussion.

Despite the fact that we did not formally elicit parameters at this stage, after the workshops it was already possible from the discussion, the literature, and our understanding to roughly indicate the expected relationships (e.g., respiratory infection is highly likely to activate the immune system) with ‘indicative’ BN parameters. These were useful for follow up discussion, and ensured that we specified the states of the variables properly and consolidated our understanding. We also identified some typical clinical scenarios (e.g., common ways the disease progresses).

We followed up with one-on-one sessions as needed, either with the same experts or supplementary experts with additional expertise. The goals were to clarify and refine the causal structure by discussing it directly, but also to perform a particular kind of validation exercise: checking that the proposed structure with indicative parameters accounted for the typical scenarios.

We then presented the model to independent experts in one-on-one sessions. The stated goals were the same as the previous sessions, but if the new experts’ answers did not substantially differ from those of the previous experts, then this also provided a more thorough form of validation.

Finally, we sent the refined models to all the participating experts for inspection before publication. The goals were to verify widespread endorsement as a broader form of validation, and also to show participants that their efforts had yielded (if endorsed) a finished product.

Overall, 35 different experts contributed a total of 126 h of face-to-face time to the group workshops and one-on-one meetings, with some additional activities also assisting in model development. A more detailed quantitative breakdown is as follows:

- 2 subject-matter surveys were sent out prior to early group workshops, which were answered by approximately 55 different experts.
- 7 group workshops were held in which 26 different experts participated (some attending more than one), contributing a total of 106 expert hours.
- 15 one-on-one follow-up meetings were held (occasionally two experts attended a meeting) in which 15 different experts participated, contributing a total of 20 expert hours. Where one-on-one meetings focused on reviewing a model produced in a group workshop (11/15 meetings), these almost always (10/11) involved new experts who had not already participated in developing the model discussed.
- After the structures of the expert BNs had been determined, there were subsequent meetings (11 meetings involving 35 different experts contributing a total of 85 h) focused mainly on parameterization of our *Progression* model via available databases, which provided as a byproduct some additional degree of detail and validation for the experts’ theoretical causal BNs presented here, e.g., for the definitions of some of the variable states.

Fig. 4 illustrates the same sequence of elicitation session types (left column), and how they relate to two more detailed elicitation procedures. To maximize the value of each session, a sequence of associated tasks was performed before and after the session itself (middle column). This work was most extensive for the group elicitation session, and is detailed below, but the work was repeated to a lesser extent for each of the one-on-one sessions.

**Fig 4.**
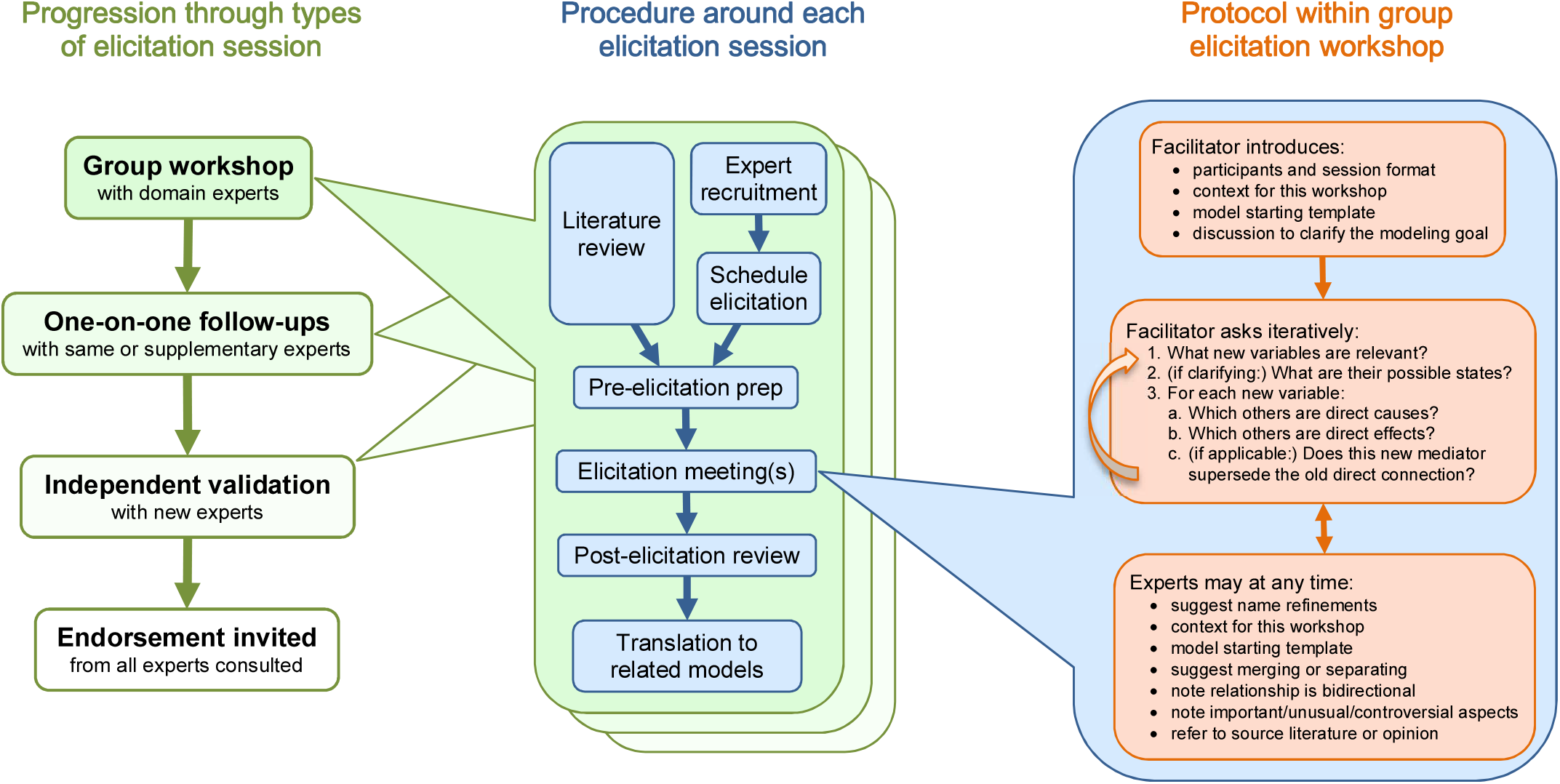
Nested BN elicitation procedures. Overall, we proceeded from group workshops to final endorsement (left column). For each type of session, several tasks were performed before and after it (middle column), with more work usually required for the earlier types of session. Within the group workshops, an efficient cooperative protocol was used consistently, encompassing introductions, stimulus questions, and ad hoc responses (right column).

To conduct a group session, we followed a particular elicitation procedure (right column) guiding the introduction, efficient elicitation questions for causal structure, and the kinds of interjections from experts we encouraged. In contrast, for each one-on-one session, a customized agenda was followed with the issues and scenarios to address via more flexible interactions.

### 2.4 Procedure around group workshop

#### 1. Literature review

Coordinators searched for and reviewed relevant articles in the literature. They identified areas of expertise required and some controversial issues. This work continued in parallel to the next two steps.

#### 2. Expert survey and recruitment

TS leveraged existing clinical and research networks to enlist contributions from domain experts. We directly emailed individuals as well as a national infectious diseases clinical mailing list, inviting them to complete a web-based survey (via the Survey Monkey tool). This served multiple purposes. First, it elicited initial information on the potential causal model. Since we expected many people would not fully complete the survey, we shuffled blocks of questions randomly so that we obtained a similar number of responses to all questions. Second, at the end of the survey, we asked if respondents wished to participate further in group workshops, so it functioned as a preparatory exercise for any participants of the workshop. Third, since it was a long survey (100 questions over 5 pages), the amount participants managed to complete was taken to be an implicit measure of their engagement. We then used other techniques to increase engagement before the workshop, such as distributing recent model diagrams and descriptions (or important excerpts), and asking a few more open-ended questions.

#### 3. Workshop scheduling

Each week, we collectively decided which workshop to schedule next by reviewing our progress and identifying the most pressing need. We invited suitable experts with limited notice (2 weeks) but achieved good participation rates, which we attribute to a strong motivation to contribute to the pandemic response as well as, in some cases, existing relationships with our team. Since not everyone who was willing to contribute could be available at any given time, we invited more people than we could optimally accommodate, and anyone who could not attend was invited to validate the model in a subsequent round. We scheduled 2 h timeslots, since in our experience this is the minimum to introduce experts to the technology and to the problem and still have adequate time to elicit their opinions. This is also the maximum time that most of our experts could contribute (due to limited time or attention), and most indicated a preference for shorter sessions. We tried to run the sessions as efficiently as possible, but used the full 2 h in every case.

#### 4. Pre-workshop preparation

Prior to each workshop, we gave participants background material that identified issues and summarized features of the model to be discussed and developed during the workshop. We used the literature review to prepare a starting template for the model. This gave our experts something to critique and expand upon, rather than starting from a blank slate, and hence expedited discussion and elicitation. We also presented a schematic overview of how each submodel would fit in with the others, and an explanation of the goals of our project.

#### 5. Elicitation workshop

We began each workshop with a short presentation that included “housekeeping” such as introductions and the format of the session, a summary of the problem space and the goals of the workshop, the starting template for the model, and any other supporting materials. We raised the general issue of what question the model is trying to answer, and tried to achieve an expert consensus on this. We then proceeded to a standard set of questions for eliciting causal structure, applied in an adaptive and iterative fashion. We asked about which factors (variables) are relevant, and only asked about their possible states if it was important to help define them. Experts frequently suggested clarification of the concepts intended to be captured by existing variables or modification of their names as elicitation progressed, and sometimes suggested merging and separating variables. We asked about the causal relationships between them, usually focusing on one variable at a time and asking which other variables are direct causes and which are direct effects (a localized version of the “bow tie” method used for risk analysis [49]). When new variables were added and connected, we also asked if this superseded any prior direct connection between a parent and a child of the new variable. Experts sometimes pointed out that a relationship could be bidirectional, which we noted, but in this initial non-dynamic graph we displayed only one direction. We chose a direction that created no cycle, and was either the initial influence, or the more immediate (i.e., with less time delay), or the strongest, as deemed most informative for disease progression or most pertinent to a submodel’s goal. We also noted any other comments, such as which connections were more important, unusual, or controversial. Experts sometimes clarified the basis for their suggestions by referring to the literature or their own opinion. The primary outcome of the workshop was a causal DAG (in the GeNIe BN software) that a high proportion of experts agreed captured the most important underlying pathophysiological processes.

#### 6. Post-workshop review

After the workshop, our team had a debrief discussion (30**–**60 min), for feedback on their own performance and to consolidate their understanding. Subsequently, the modelers cleaned up any loose ends in the model or documentation, making sure all the knowledge elicited was understood and documented correctly, and simultaneously looking for gaps that would need further clarification. This included indicative parameters and typical scenarios, as described above. The whole workshop, conducted remotely by web-based videoconference (Zoom), was recorded as video, and the audio later transcribed into text. This allowed us to review any part of the elicitation as required. Our transcripts and video recordings, the “raw data” of these elicitation sessions, have been retained and securely stored. We used our newfound understanding to choose and schedule subsequent elicitations and actions. These included emailing questions to specific experts that were present or to other experts with different skill sets or knowledge, and then if needed, scheduling one-on-one elicitation sessions.

#### 7. Translation to application models

As a result of our various expert elicitation sessions, we produced multiple “expert models” that represented different body subsystems, and sometimes represented alternative perspectives put forward by different experts. Our approach here was exploratory, avoiding premature judgments on what would be important for practical applications, so we tried to capture this theoretical expert knowledge comprehensively. However, our ultimate aim is to develop practical tools powered by customized “application models”, the first of which will be to support clinical reasoning in the prognostication for COVID-19. The expert models will provide a theoretical rationale for, and increase confidence in, the validity of the application models. Our modelers decide which aspects of the expert models are most relevant and should be included in an application model, given its purpose and required outcomes. In some respects an application model may be simplified, e.g., by combining several latent variables. In other respects, it may be more sophisticated, e.g., by creating a DBN representing the whole system based on multiple expert BNs representing subsystems. The application models we have already developed are currently being independently validated by experts, both structurally and quantitatively, as well as validated by their fit to the data.

### 2.5 Adaptive, spiral processes

Although we depict our methods as several linear sequences, our description should make clear that in combination these formed a spiral process, as widely recommended for model building: i.e., our sequences of steps were iterated to produce successive improvement. Most obviously, the sequence depicted in Fig. 3 is designed to successively refine and validate elements of the model through repeated elicitation.

In the same vein, we also used an adaptive elicitation procedure, i.e., we used the outcomes of elicitation and modeling exercises to help decide what sort of elicitation and modeling should be done next. So, to some extent, we learned and refined our methods over the course of the project.

One example of adaptive elicitation was modifying our pre-workshop material for participants. For our early workshops, we prepared surveys on issues where we identified disagreement in the literature. We aimed to engage and prepare participants, and obtain a preliminary understanding of their views. However, despite the assistance of our coordinators, we found it difficult to design survey questions that our modelers felt were relevant to the modeling choices, yet were framed and expressed in a way that most medical experts felt they could respond to. For example, experts often felt the answers were dependent on unstated contextual details.

For subsequent workshops, we simply gave participants background material that identified such issues of disagreement, and summarized features of the model to be discussed and developed during the workshop. In our video conferences, where elicitation and model building were more coordinated and interactive, we found that modelers and medical experts had much less difficulty in translating medical knowledge to models and vice versa.

Another example of adaptive elicitation was adding post-workshop supplementary meetings. After holding 2 h elicitation workshops with groups of medical experts to generate a consensus BN, it became clear that follow-up one-on-one meetings would be useful, with some of the workshop participants and sometimes with other experts who had different expertise. These meetings allowed experts who wanted additional time to give more detailed feedback on the model, and they were also a better forum for resolving persistent points of disagreement or uncertainty.

## 3 Results and Discussion

### 3.1 Models produced

Fig. 5 illustrates the history of our model development in a cladogram. The four models we commenced with, as described in § 2, did not survive in their initial forms. As an adaptive, generative process in a complex, novel domain, our progress was inevitably evolutionary: new model variants were created, crossbred and adapted for each environmental niche, with many now extinct and a few still extant.

**Fig 5.**
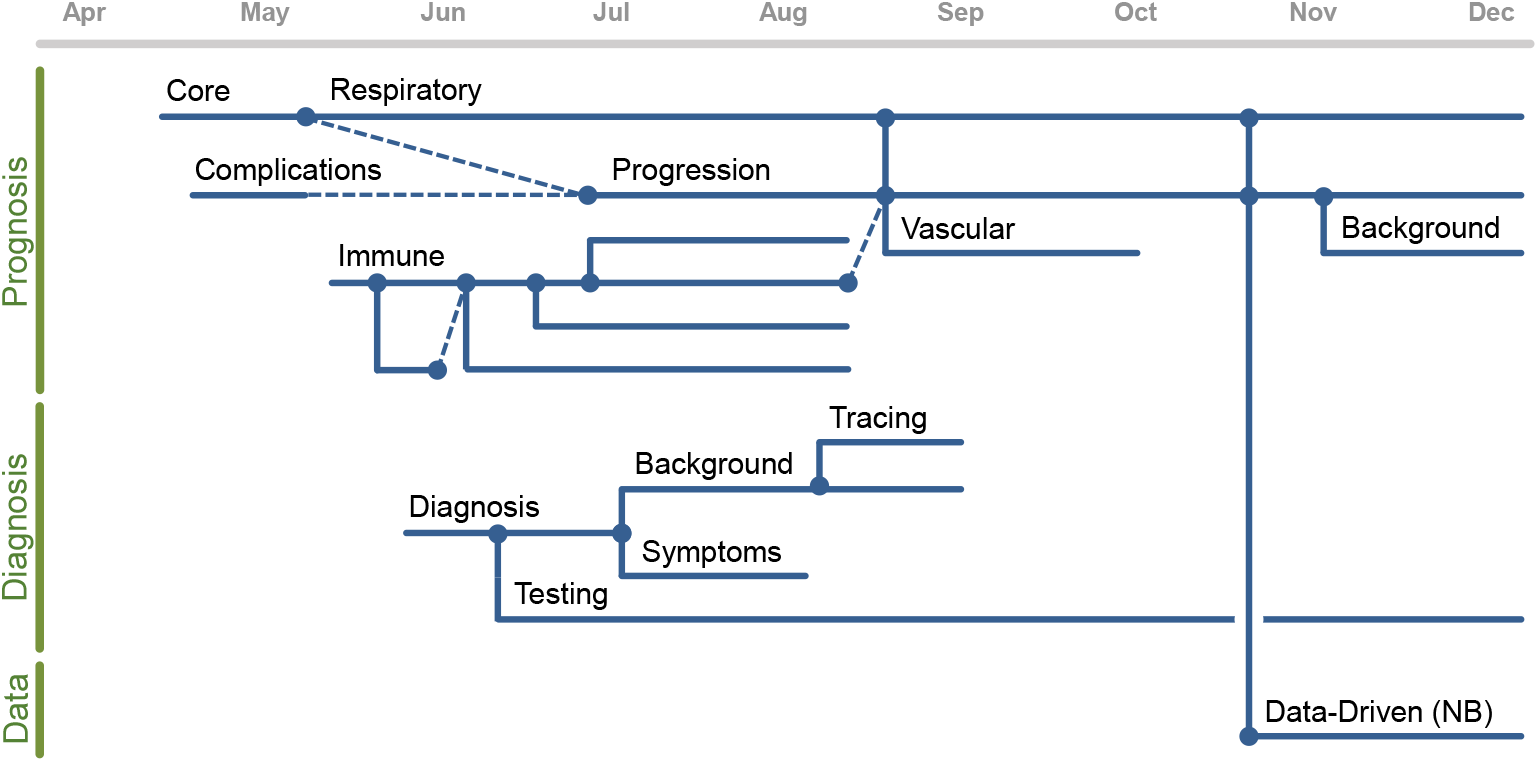
Model cladogram from April to December 2020. Some model lines primarily concerned prognosis (top family), other lines primarily concerned diagnosis (middle), and one line concerns Naïve Bayes (NB) and related variants adapted for preliminary parameterization and data exploration using available data (bottom).

Our *Core Mechanism* model centered mainly on the respiratory system from the outset; as the multi-system nature of COVID-19 became apparent we decided to rename it the *Respiratory* model and focus on the initial infection process in that system. It is presented below.

Our *Complications* model largely begins where the *Respiratory* model’s story of pathophysiology ends, capturing for more severe cases the knock-on effects to other organ systems and body functions. After a period of work on both this and the *Respiratory* model, key ideas were identified from both that would form the basis of an application model, called the *Progression* model.

This *Progression* model subsequently came to incorporate ideas from further expert models, including the *Immune System, Vascular System* and *Background* models, and it became clear that this BN would need to be transformed into a DBN, which can better represent longer-term system dynamics by replicating variables at different time steps (§ 1.1), in order to track the longer-term progression of COVID-19 and ultimately assist with prognosis. The parameterized *Progression* model changes shape depending on the data set and setting, so it is not presented here; we are currently preparing the detailed presentation it requires. However, the expert *Complications* model on which it is most closely based is presented below.

We invested considerable effort in modeling the *Immune System* response, which proved to be a complex and contentious topic with much uncertainty and little that could be confidently extrapolated from other infections. However, after this investigation we concluded that much of the exact detail of the mediators of the immune response might be unnecessary for the purposes of diagnosis and prognosis where we ultimately aimed to contribute practical tools. Hence, we deferred work on this model and it is not presented here.

After some work eliciting details for our *Diagnosis* model, we made the judgment that with the emergence of sensitive and specific diagnostics, diagnosis of patients was not a task for which our models were likely to provide much assistance, with the exception of the specific task of managing COVID-19 testing in the commu-nity. We therefore discontinued modeling most of these details, and continued modeling the remainder in our *Testing* model. We have already published this as a causal BN [50], and will not reproduce it here.

We detail these procedural missteps and ‘dead-ends’ to accurately describe a method of modeling, whereas omitting or rewriting this history might give the misleading impression that the two models presented below were always exactly the intended output.

### 3.2 Method of presentation

Since COVID-19 pathophysiology is complex, we developed several BNs for different subsystems; here we present our two main products. Our *Respiratory* BN concerns the detailed pathways that link the initiating infection to respiratory pathophysiology and the beginning of complications, and hence it focuses primarily on the lungs. Our *Complications* BN concerns higher-level interactions and how the disease may progress after complications have begun, and hence it includes other organs that become important later in the progression of a severe case.

We present the models in three complementary types of file:

1. **Causal DAGs and BNs**, in which we specify the causal structures in supplementary static graph diagrams (S1 Fig., S2 Fig.) and in supplementary GeNIe BN files (S1 Model, S2 Model). GeNIe can present the structures simply as causal DAGs, or more dynamically: variables can be displayed with their states and indicative probabilities, and the states of variables can be specified to explore how this changes the probability distributions over other variables’ states.
2. **BN descriptions and issues**, which summarize the processes these graphs represent (§ 3.4 and 3.6), and discuss a few unusual or controversial features (§ 3.5 and 3.7).
3. **BN dictionaries with references**, which give the definitions of variables and states, and also cite some of the research literature that supported specific BN features. We present illustrative examples in Fig. 2, discuss some general features in § 3.8, and provide the complete documents as supplementary files (S1 Table, S2 Table).

### 3.3 Limitations of our models

While there are still unknowns and competing hypotheses in the literature, the differences are not profound enough to require presentation as competing DAGs; our list of issues is sufficient to indicate which structural components would differ. Some disagreements only concern the degree of impact of alternative pathways, which corresponds to differing numerical parameterizations of the BNs, and no parameterizations are included here.

Both our BNs do not include important background factors such as age, comorbidities and vaccination status, that strongly influence the probability of more serious COVID-19 outcomes (and each other). Knowledge about their role in the COVID-19 process was too limited: although they are known to directly influence some of our variables (e.g., vaccination reduces the chance of initial infection), not all of their direct influences are clear (e.g., vaccination also decreases the chance of infection developing into severe COVID-19, probably by influencing multiple variables along these pathways). Fortunately, these theoretical models are useful and valid without them. According to our experts’ assessment, the distinctive COVID-19 causal structure depicted is unlikely to change for any particular specification of background factors. Rather, such factors will affect the parameters, e.g., how strongly some variables influence others. When adequate data is available to adjust these parameters depending on background factors, then these factors can be appended by researchers to the DAGs we present here as additional causes.^3^

Many variables involved in these processes are latent, but their probability distributions can be inferred from observed evidence, such as clinical signs, symptoms and laboratory measurements. Again, our BNs do not include all the relevant possible evidence available now or in the future, but our model is valid without them. Structurally, they can be appended as additional effects of some of the variables in our DAGs, without changing the existing DAG connections. As data becomes available (e.g., the false positive and false negative rates for a new test) then their CPT parameters can be estimated.

Feedback loops, i.e., variables that influence each other over time, are not included in these acyclic models. Some are, however, noted in the BN dictionary. Feedback loops, like background variables, are explicitly included in our parameterized *Progression* DBN for modeling longer-term COVID-19 trajectories. The details will be presented in a future publication on this model and associated practical tools.

### 3.4 *Respiratory* BN Description

Our *Respiratory* causal DAG (S1 Fig.) models the physiological process underlying COVID-19 in the respiratory system, outlining multiple and often concurrent pathways from the initial replication of the virus to key downstream complications such as multi-organ failure.

We color-coded the nodes: pink for the **Infection** process, yellow for more detailed mechanisms relevant to the **Pulmonary** system, orange for those **Complications** that mainly arise directly from the respiratory system, and cyan for a selection of **Signs and Symptoms** included only for illustrative purposes. “Virus” refers to SARS-CoV-2.

References have been omitted from the following description, but can be found in the associated variable entries in the BN dictionary.

#### Infection process

After an exposure event, the virus can enter the upper respiratory tract via the nasopharynx (Virus enters upper respiratory tract) and may replicate locally and cause inflammation at various locations in the respiratory system, including the epithelial cells of the upper respiratory tract (Upper respiratory tract epithelial infection) and the lower respiratory tract (Alveolar epithelial infection). Direct infection involving the alveoli without preceding or simultaneous infection involving the upper respiratory tract is possible, although it may be less likely.

The virus can first become established at one site then spread to or re-establish in another. For example, Infection of olfactory epithelium following epithelial infection in the upper respiratory tract has manifested as Ageusia and/or anosmia in some patients. The virus may spread from epithelial to endothelial cells in alveoli, causing Alveolar endothelial infection. During all infection processes, the virus may enter the bloodstream (Viremia) and potentially spread to other parts of the body (not included here).

Viral replication at any site may activate Systemic immune and inflammatory response, which often results in the release of pro- and/or anti-inflammatory blood markers from immune-related cells.

#### Pulmonary system

The primary function of the respiratory system is to support the metabolic processes of the body by taking in oxygen and removing carbon dioxide; this occurs through gas exchange between air and blood at the delicate membranes of the lung alveoli. Our *Respiratory* causal DAG (S1 Fig.) models the initial pathophysiological process which is initiated by SARS-CoV-2 infection in this system.

The most notable large-scale feature is the three distinct pathways from the initial infection to involvement of the respiratory system and then complications, although these pathways have interconnections and often contribute concurrently.

#### Mechanical pathway

Alveolar and systemic inflammation can damage pulmonary capillaries causing the leakage of plasma (Pulmonary capillary leakage), leading to filling of the alveoli with exudate (Alveolar consolidation) that causes stiffening and reduced mechanical lung function (Reduced lung compliance), and results in Ventilatory insufficiency. Alveolar epithelial infection (and inflammation) can also directly reduce compliance, but to a lesser extent. Another possible cause of ventilatory insufficiency is a loss of muscle mass (Muscle wasting) which can be directly caused by catabolism as a result of the systemic immune response and may be affected by a range of background factors, such as nutritional status and immobility, not shown in this model.

#### Gas exchange pathway

Pulmonary capillary leakage and alveolar consolidation blocks the passage of air and reduces the alveolar surface area available for gas exchange, and this mismatch between ventilation and blood perfusion reduces gas exchange in the affected parts of the lungs (V/Q mismatch). At an extreme, local perfusion may occur in the absence of ventilation with oxygenated air (Shunt). Alveolar infection causes local Hypoxia which can trigger counteracting Pulmonary vasoconstriction, an adaptive physiological response which helps to match regional perfusion to ventilation in the lungs and thus reduces the extent of V/Q mismatch.

#### Coagulation pathway

Endothelial infection/ inflammation and possibly the presence of virus in the blood (viremia) may trigger a systemic immune and inflammatory response, and this may result in an abnormally high propensity to coagulation (Hypercoagulable state). This may begin locally in the small vessels of the lower airways (Pulmonary microthromboses), but can later manifest as macroscopic thromboses in larger vessels (Other thromboses). Both types of thrombosis can block vessels in the lungs (Pulmonary circulatory blockage) and lead to V/Q mismatch, to which alveolar endothelial infection/inflammation can also directly contribute.

The direct consequences of impaired lung function, due to any of these pathways, include insufficient blood oxygenation (Hypoxemia) and excessive blood carbon dioxide (Hypercapnia), which can be measured using blood gas assays or by pulse oximetry for oxygen. Furthermore, pulmonary vasoconstriction caused by hypoxia and pulmonary circulatory obstruction caused by thrombosis can produce raised blood pressure in the pulmonary arteries (Pulmonary hypertension), which interacts with the rest of the system in a complicated way that is not explicitly described here, but which can lead to Impaired cardiac output.

#### Complications

The *Respiratory* model includes and summarizes some key complications. The lungs interact most closely with the heart; so in addition to pulmonary hypertension, pulmonary thrombosis can directly cause impaired cardiac output. Systemic immune response and insufficient oxygen in the blood (i.e., hypoxemia) can also lead to impaired cardiac output via Diminished myocardial contractility.

Maintaining end organ tissue oxygenation relies on sufficient oxygen transport via the blood, so Hypoxia occurs where there is hypoxemia or insufficient blood perfusion due to impaired cardiac output. Hypoxia can directly cause organ failure, or Multi-organ failure if more than one is affected (which is often the case).

Other pathways that can eventually lead to multi-organ failure include virus in the blood-stream causing direct viral injury to organs, and an extreme systemic immune response (i.e., cytokine storm).

Several mechanisms can trigger the body’s sensor for insufficient oxygen, including reduced compliance (i.e., a mechanical failure of the lungs), hypoxemia, hypercapnia, and acidosis (both respiratory and metabolic). If the body senses there is insufficient oxygen supply (Perceived need for air), it demands more oxygen intake, which manifests as Dyspnea, one of the most important symptoms for COVID-19.

### 3.5 *Respiratory* BN Issues

We describe here four issues with our *Respiratory* model that seem noteworthy, either in themselves or as illustrative examples. Some issues involve modeling choices: even where the process being represented is entirely agreed upon, modeling sometimes involves choices about how to represent it, and some of the options we chose here may need clarification or appear controversial. Some issues involve domain knowledge: there are some aspects of COVID-19 that are still not fully understood, about which there may be competing hypotheses. So, some of the options we chose here may need revision in future as further research resolves these controversies.

#### Upper respiratory tract mediation

One theoretical controversy, related to the diagnostic value of a dry cough, concerns three possible respiratory pathways by which the virus enters the body. Respiratory viruses are thought to typically infect the upper respiratory tract first (abbreviated as URT in the BN), then descend by contiguous spread to the alveoli and (rarely) via the bloodstream. COVID-19 may take this normal path, or may bypass the upper respiratory tract or the larger airways to infect the endothelium of the lower airways more directly via the ACE2 receptor, or less directly from the upper respiratory tract via the bloodstream. The data on this issue is currently limited so we have included all three pathways in the model, but our experts thought the typical path for respiratory viruses was less likely than the other two. Future findings may support this opinion and clarify their relative importance.

#### Vascular lung damage feedback loops

A high degree of vascular lung damage, caused by microthromboses in lung capillaries, appears to be a unique feature of COVID-19. Our *Respiratory* BN includes vascular involvement (in the **Coagulation** pathway) primarily as a mediator for the initial phase of infection in the lung, whereas our *Complications* BN places more emphasis on subsequent feedback loops. Since vascular involvement is complicated, we explored it by eliciting a separate vascular model from two of our experts, which is not presented here. We were satisfied that the vascular elements included in our *Respiratory* and *Complications* models were sufficient for our purposes.

#### V/Q mismatch and Shunt seem logically related

It is good practice to avoid including logically related terms in a causal model, i.e., where their semantics results in a logical dependence. Such a connection results in probabilistic dependence and an arc in the model, but it may behave differently to a causal connection under intervention, which can make it difficult to adapt the model for decision support. Here we included both Shunt and V/Q mismatch, even though shunt is (by definition) a type of V/Q mismatch, so the former logically entails the latter. This accurately represents the statements elicited: our experts consistently used both these terms and distinguished between them. They had good reason: different interventions (such as positive airway pressure for shunt and supplementary oxygen for V/Q mismatch) are considered appropriate in each case. So, mirroring their terminology faithfully captures expert knowledge in a way they can agree is accurate, which in this case includes the relevant causal connections to interventions or tests.

#### Neurological explanation for “happy hypoxics”

We model the patient’s perceived need for air separately from dyspnea (shortness of breath). An unusual feature of COVID has been patients who present as “happy hypoxics” (physiologically hypoxic but without symptoms) [51]. One causal hypothesis is that neurological viral infection inhibits the perceived need for air. However, at the time of modeling this connection was very speculative, so we decided not to include this pathway, pending further medical research.

### 3.6 *Complications* BN description

Our *Complications* causal DAG (S2 Fig.) models the main physiological processes underlying the progression of COVID-19 from the initial infection in the respiratory system to complications in other organs. As before, the BN dictionary notes some key feedback loops and provides references for the mechanisms associated with each variable mentioned below.

#### Overview

The model explicitly describes the status (dysfunction) of nine key organ systems, namely, respiratory, vascular, cardiac, liver, kidney, hematologic, gastrointestinal, cortical and brainstem dysfunctions. Among these, we choose to paint a more fine-grained picture of the **Vascular** (nodes in blue) and **Cardiac** (nodes in green) systems due to their potential earlier involvement, as well as more significant **System-wide** impact (nodes in off-white) on **Other organs** (nodes in orange).

The full picture of the pulmonary system is shown in the *Respiratory* model; here we retained a few key **Pulmonary** variables (nodes in yellow) and two major consequences of pulmonary dysfunction due to their system-wide impact: Hypoxemia and Hypercapnia. Two **Background** factors (nodes in purple) are added only for illustrative purposes.

#### Pulmonary system and direct viral impact

Despite its initial establishment in the pulmonary system, infection with SARS-CoV-2 can potentially drive the dysfunction of all listed organ systems directly via two main mechanisms, Direct viral injury and the Systemic immune/inflammatory response, throughout the entire process of the disease. In particular, they both can affect the vascular system by reducing Vascular integrity and Vascular tone. Vascular integrity refers to the status of endothelial structure compromising permeability, and the vascular tone refers to the degree of constriction of a blood vessel.

The systemic inflammatory response can additionally increase the probability of Hypercoagulable state (as also depicted in the *Respiratory* model) and Dehydration. For the cardiac system, direct viral injury and the systemic inflammatory response might both result in Acute cardiac inflammation.

#### Vascular system

The vascular function (in this model) represents the ability of blood vessels to ensure sufficient blood circulation to meet the metabolic needs and energy requirements of the organs. Reduced vascular tone, Reduced vascular integrity and Hypercoagulable state are three major mechanisms that contribute to vascular dysfunction.

The first two mechanisms can both lead to Reduced functional intravascular volume, i.e., less blood is available to supply the organs because of the loss of vascular tone or the shift of intravascular fluid to the interstitium (Fluid shift to interstitium) due to altered permeability (vascular integrity).

The hypercoagulable state may increase the vascular resistance to flow of blood through affected vessels. Abnormally low vascular resistance is also problematic because it can lead to insufficient pressures needed to ensure the distribution of blood and perfusion of the organs.

#### Cardiac system incl. Vascular interaction

The cardiac and vascular systems closely interact. While vascular resistance and functional intravascular volume predominately drive the amount of blood available to supply organs, the heart creates the forward movement of blood needed to maintain supply to organs (organ perfusion). This is measured as Stroke volume (the volume of blood ejected for each stroke) and Cardiac output (the product of heart rate, not shown, and stroke volume).

Acute cardiac inflammation and Ischemic cardiac injury can both cause abrupt or gradual deterioration in the cardiac output, either by inducing Abnormal contractility of the heart (the strength with which it pumps), or the synchronicity and efficiency with which it pumps; the latter can be manifest as abnormal heart rate or arrhythmia.

Hypercoagulable state can lead to insufficient supply of blood more directly and acutely by blocking the pathway to certain parts of body; this can cause ischemic cardiac injury and pulmonary hypertension in the heart and lungs, respectively.

#### Other organ dysfunctions and failures

The ultimate need of organs is enough oxygen and metabolites supplied through the blood circulation to maintain their vital functions, we have therefore replaced Hypoxia from the *Respiratory* model with separate nodes that distinguish between the supply of blood (as organ perfusion) and the supply of oxygen and metabolites. Reduced supply of blood will lead to reduced supply of oxygen and metabolites, and the latter will also be reduced if there is a lack of oxygen in blood (hypoxemia) even if the supply of blood is normal.

Reduced supply of oxygen and metabolites can lead to dysfunction of any of the listed end-organs, which often further disrupts the balance of the whole system. For example, liver, kidney and gut play important roles in maintaining the balance of electrolytes, metabolites and acids. The dysfunction of these organs would generate feedback loops with other organs including pulmonary, cardiac, and vascular systems via electrolyte and metabolite imbalance and acidosis. In this model, we summarize all such feedback loops by presenting only their final deterioration to particular organ failures.

Since vascular failure is ill defined, we instead represent a key pathway in which a hypercoagulable state combined with possible liver, kidney and hematologic dysfunctions lead to general coagulopathy that can in turn contribute to severe vascular dysfunction that may result in the heart stopping and subsequent death. Critical failures of the brainstem, pulmonary, and cardiac systems also rapidly lead to an inability to support any of the vital organ functions and are terminal events.

### 3.7 *Complications* BN issues

As we did for the *Respiratory* model, here we describe two issues with our *Complications* model that seem noteworthy, involving modeling choices and/or domain knowledge.

#### Hypercoagulable state and Coagulability seem logically related

The model includes the Boolean variable Hypercoagulable state, and also a more general variable Coagulability, which has states to represent both hypercoagulability and hypocoagulability. This reflects the way our experts consistently preferred to describe the system, and our primary goal in this model was to capture expert knowledge in a way they can agree is accurate. It may be efficient to focus on a particular state, such as hypercoagulability, where that has a higher frequency or specific effects that other states do not. However, a reasonable alternative would be to represent Coagulability twice, at two different time steps, with hypercoagulable as one possible state of this variable—which is what we use in our *Progression* DBN model.

#### Lung–acidosis feedback loop

The *Complications* model involves several important feedback loops. For example, as noted in the description, if the lungs are damaged and do not adequately remove CO_2_, then the resulting acidosis may damage the lungs themselves.

### 3.8 BN Dictionaries with References

All variables in these two BNs have both descriptions and their relationships with their parent nodes (supported by selected references) specified in their Dictionaries (S1 Table, S2 Table); a few illustrative examples are given in Fig. 2. Here we discuss some general features.

#### Definitions and databases

Chronic pulmonary disease in Fig. 2 illustrates that many of our variable descriptions are explicitly aligned with the case database, e.g., IDDO (https://www.iddo.org/covid-19) or LEOSS (https://leoss.net/), that we will use to parameterize both our expert and application BN models. If other researchers use slightly different descriptions and datasets, these are unlikely to alter the relevant causal connections, though different datasets in particular may not be sufficiently detailed to support each relation and, as is always the case, will need appropriate processing.

#### Observable versus latent

Symptoms such as Ageusia and/or anosmia or test results such as Low oxygen saturation (SaO_2_) are classified as directly observable (purple), and can often be represented as a node influenced by (and considered a noisy measurement of) a few underlying variables, but does not itself influence those underlying processes. Hence, additional observations can usually be represented simply by adding another such variable without changing the underlying causal structure. For simplicity, we omit these variables here, with the exception of Low oxygen saturation (SaO_2_) in the *Respiratory* model. Most other variables are classified as latent, even if, like Hypoxemia, their state can be easily and reliably determined via an observation like the SaO_2_ test.

#### Number of variable states

Many of our variables, such as Ageusia and/or anosmia, are Boolean: their only possible states are true or false. The names of these variables generally refer to the abnormal status of a mechanism, since people more readily interpret abnormal states as causes, particularly when they tend to produce an abnormal result [52], and to make the causal diagrams more easily digested in the absence of visible states. Most of our other variables, such as Acidosis, are ordered categorical: they have a finite number of discrete states in a meaningful order (usually degrees of severity). Continuous variables are often discretized during a measurement process and/or during BN construction to assist with probability estimation and computation (although BNs can include them). The states are not depicted in the causal DAG, but may be inspected in the BN GeNIe file.

#### Minimizing the state space

Many variables were mentioned during elicitation and considered as candidates for inclusion, but ultimately excluded from the BN as unnecessary complications that would only make the BN harder to understand and parameterize. Several such examples exist: Electrolyte imbalance was soon replaced with the more specific Acidosis; Acute cardiac injury and Type I/II myocardial infarction were exchanged in preference for the more general Ischemic cardiac injury and Acute cardiac inflammation, along with other associated changes; several concepts surrounding innate versus adaptive immunity and finer distinctions between immune and inflammatory response were shelved in favor of a simple node Systemic immune/inflam. response representing the overall severity of the response. Similarly, we have included sufficient states to adequately represent the elicited causal relationships; adding further states would not change the causal structure significantly, but would add superfluous complexity. For example, in the *Respiratory* model, Hypercapnia and Acidosis are represented with just two states, while Hypoxemia and Low oxygen saturation are represented with three, due to the conclusions of that model being particularly sensitive to the latter nodes.

#### Qualitative vs. quantitative influence

In principle, the relationships depicted in BNs can be highly complex, so a causal DAG alone could be hard to interpret without examining the details of the parameters. In practical applications such as this one, however, the reader usually has some basic domain knowledge which can be augmented by brief descriptions such as the ones we have given for the BN as a whole, and in the Dictionaries for each variable’s parents. It then becomes fairly easy to interpret the kind of influence indicated by each of the arcs. For example, Upper respiratory tract epithelial infection is shown as a parent of Alveolar epithelial infection, which does not in itself indicate how the parent affects the child (unless additional markup is applied, such as plus and minus symbols), but the natural and correct reading here is that the former increases the probability of the latter. Our BN files do include parameters that roughly indicate a degree and direction of influence. These were often suggested by our experts during structure elicitation, and used to assist structure validation, but they were not formally elicited or estimated. Precise effects may vary with the local context of application, and precise parameters are often learned from an appropriate local dataset.

## 4 Conclusion

### 4.1 Value of our method

Methodologically, we have demonstrated how to implement best practice recommendations in a systematic, expanded procedure for developing BNs via expert elicitation. This can be emulated, adapted and refined in future projects. It is particularly well suited to projects modeling emergent diseases, since (i) it uses large-scale online expert elicitation to develop causal structure, rather than only using data or literature directly, and (ii) it produces a family of well-documented theoretical models that make the expert knowledge freely available and that can be updated as the knowledge base develops, rather than only producing a few practical BN tools that can be derived from, and supported by, the hidden theoretical base.

#### Harnesses worldwide expertise when data and literature are lacking

We demonstrated how to overcome the scarcity of data, and the exceptional abundance and unreliability of research literature, by using groups of volunteer, independent, specialist domain experts to filter, interpret and discuss the literature findings and develop a reasonable current consensus. We also utilized intermediary experts to facilitate the exchange of knowledge from domain specialists to BN modelers and vice versa.

This approach is unusual, and the total number of expert hours required was exceptionally high. We obtained these hours by recruiting a large number of experts to contribute the time they could spare, which was facilitated by our extensive use of online meetings. In addition to being COVID-safe, they had the logistical advantages that experts could meet briefly within their busy schedules from their normal places of work and in highly dispersed locations, without the overhead of travel time or even needing to be in the same country, which effectively broadened our pool of available experts and the number of elicitation sessions which were feasible. Online tools also enabled all our elicitation exchanges to be easily recorded.

#### Adaptive evolution of theoretical models that can be updated and used by others

We followed an unusual two-phase process of BN development and publication. We aimed to first elicit relatively detailed causal models that captured the experts’ understanding of the relevant processes, including theoretically salient latent variables that might not be useful in the specific, practical BN tools we aimed to subsequently develop. Furthermore, we are publishing here the full details of, and documentation for, these causal models. The goal is to make the elicited expert knowledge freely available to others in a form that can be readily updated as medical science progresses, and used for a variety of purposes.

Since both the method and domain were large, complex and novel, it was necessary to subdivide, adapt and evolve both the elicitation procedure and the models. We developed a systematic approach of refining and checking the group output with one-on-one follow-up meetings as necessary, with both group members and new, independent recruits. By enlisting new domain experts, we progressively validated and refined our models in the iterative, incremental style that is broadly recommended for BN building, but is more logistically challenging and has rarely been achieved when using expert elicitation.

Although we have specified our method in clear logical sequences, we stress that in model building, teams need to be flexible and in some respects progress is unpredictable. Pre-workshop questionnaires, which we abandoned, may prove more suited to other topics, especially where variables are known or easily identified beforehand; an elicitation workshop may reveal that substantial supplementary expertise or literature is required; it may become clear that models should be split or merged; and even after experts have signed off on a model it may need to be abandoned if subsequent testing with numerical datasets doesn’t support it.

There is also scope to refine our methods further and test some variations in future applications. For example, group workshops followed by one-on-one follow ups worked well, bringing key information and model structures together quickly (via the workshops) and then allowing more cautious validation and refinement of those structures in detail (via the follow ups). However, we now believe that conducting some one-on-one interviews may be useful earlier in the elicitation process. They may be useful prior to questionnaires to increase the chance that questions handle problematic concepts well, or prior to group workshops to provide better structure for those sessions.

### 4.2 Value of our models

#### Representing current and future understanding

The two fully documented models presented here are the first published causal DAGs of COVID-19 pathophysiology. Confidence in their quality is underwritten by the rigorous process used to create them. They can be used as a visual aid to understanding and explaining the internal causal processes and remaining controversies of COVID-19, and are accessible to audiences without specialist medical expertise.

Medical science is constantly progressing, so our models must be refined accordingly—possibly using the method we have provided here, whether employed by us or by others. Fortunately, most of our work will not need to be repeated, since elicited causal DAGs can be readily updated (§ 1.1). This includes resolutions to the theoretical issues mentioned in § 3.5 and § 3.7, as well as adding new background factors, new diagnostic tests, and new interventions.

Feedback loops are not incorporated in these DAGs, even though some are important for modeling the progression of the disease over longer time scales. We noted some of these loops in the Dictionaries, and will discuss them in future work, in conjunction with presenting our DBN model for disease progression.

#### Guiding design of empirical studies

Causal DAGs are now widely recommended to inform the design and analysis of health-related observational studies [53]. DAGs help researchers identify relevant factors, and more specifically, factors that are potential mediators, confounders, and sources of selection bias or measurement error. So, we anticipate that our models will serve as a guide to the design of cohort and case-control studies of the effects of various demographic and clinical risk factors, or of the effects of various interventions, on one of several clinical outcomes of interest for SARS-CoV-2 infection.

Causal DAGs have similar applications to the design of experimental studies, even where manipulation or randomization of the treatment variables reduces confounding. So, we anticipate that our models will also be useful to guide the design of clinical trials of treatments for SARS-CoV-2 infection.

Our DAGs can be used to identify important non-treatment factors that may influence and produce unwanted variation in the trial outcome, and which should therefore be considered for control by stratification or statistical adjustment (regardless of whether treatment randomization is employed) in order to increase the statistical power of the experiment. Similarly, our DAGs can be used to identify factors that might modify the effect of treatments, which should therefore be used to pre-specify patient subgroups.

By quantifying the pathways in our models that connect proximal outcomes such as hypoxia or biomarkers to key outcomes of interest such as the need for mechanical ventilation or death, it may be possible to design more statistically efficient trials that use the former as plausible surrogate endpoints for the latter. For example, we may use the degree to which a treatment reduces hypoxia and specific biomarkers in a larger group of patients as a more statistically powerful way of quantifying the expected reduction in the need for mechanical ventilation, which eventually occurs in a smaller subset of patients.

#### Developing and validating practical tools

Our models will be useful in the development and/or validation of fully parameterized practical tools for causal reasoning and decision support. In addition to using them for individual decision-making in a clinical setting, such tools can also be used as an aid to policy decisionmaking at a broader level, for example, by considering risks that apply to whole populations, or the effect of non-targeted interventions applied to readily identifiable patient subgroups.

Practical tools can be developed by any researchers directly from our models, or from future updated versions of them. This often involves identifying which evidence (observable) variables are included in the available datasets that will be used for parameterization. The theoretical causal structure of our DAGs can then be simplified or adapted to omit or merge some latent variables, while trying to retain sufficient latent structure to encode the expected eviden-tial dependencies. These application models are then parameterized using the datasets, and may be further modified depending upon their performance (e.g., using cross validation to assess their mean predictive accuracy on various measures for target variables given some evidence).

Decision and utility nodes may be added to the theoretical or application models, to allow causal BNs to formally model and predict the consequences of therapeutic interventions, which is essential for clinical reasoning and decision making. As new data accrues on the impacts of such interventions, the application models can be re-parameterized to remain accurate.

Even if an application model is developed independently of our models, the dependencies this application model encodes between variables (including interventions) can be checked against our DAGs to identify any likely discrepancies. Our models thus provide a ready-made and well grounded source of theoretical validation for other research teams using different modeling methods.

In our ongoing work with these BN models, we are already developing several of our own tools for either standalone or integrated decision support, parameterized using the IDDO subset of the ISARIC database (https://www.iddo.org/covid-19) and LEOSS databases (https://leoss.net/). We are tailoring two distinct tools based on an applied version of the models we have described here, one for in-hospital prognosis and management of COVID-19 patients and another to support resource management and monitoring for confirmed cases. For the in-hospital prognosis tool, available information about the patient can be entered into the tool by a clinician directly, or retrieved automatically from a patient’s record. The clinician will then receive information not just on the patient’s overall prognosis, but also on risks around organs, body functions and treatment requirements over the next 24 hours and the next 5 days. For resource management, the same application model can be used, but integrated into existing, centralized, government monitoring systems. The model would be used for confirmed cases being monitored at home or in non-medical facilities to quickly flag those at risk, allowing ministry staff to make better informed and more fine-grained decisions around utilization of hospital and medical resources.

## Data Availability

All data produced are available online at https://osf.io/bynr6/

## Supporting information

All the following supplementary files will remain available on our OSF project page (https://osf.io/bynr6/) in the versions referred to here, accompanied by any subsequent versions of them that we develop.

**S1 Fig.**
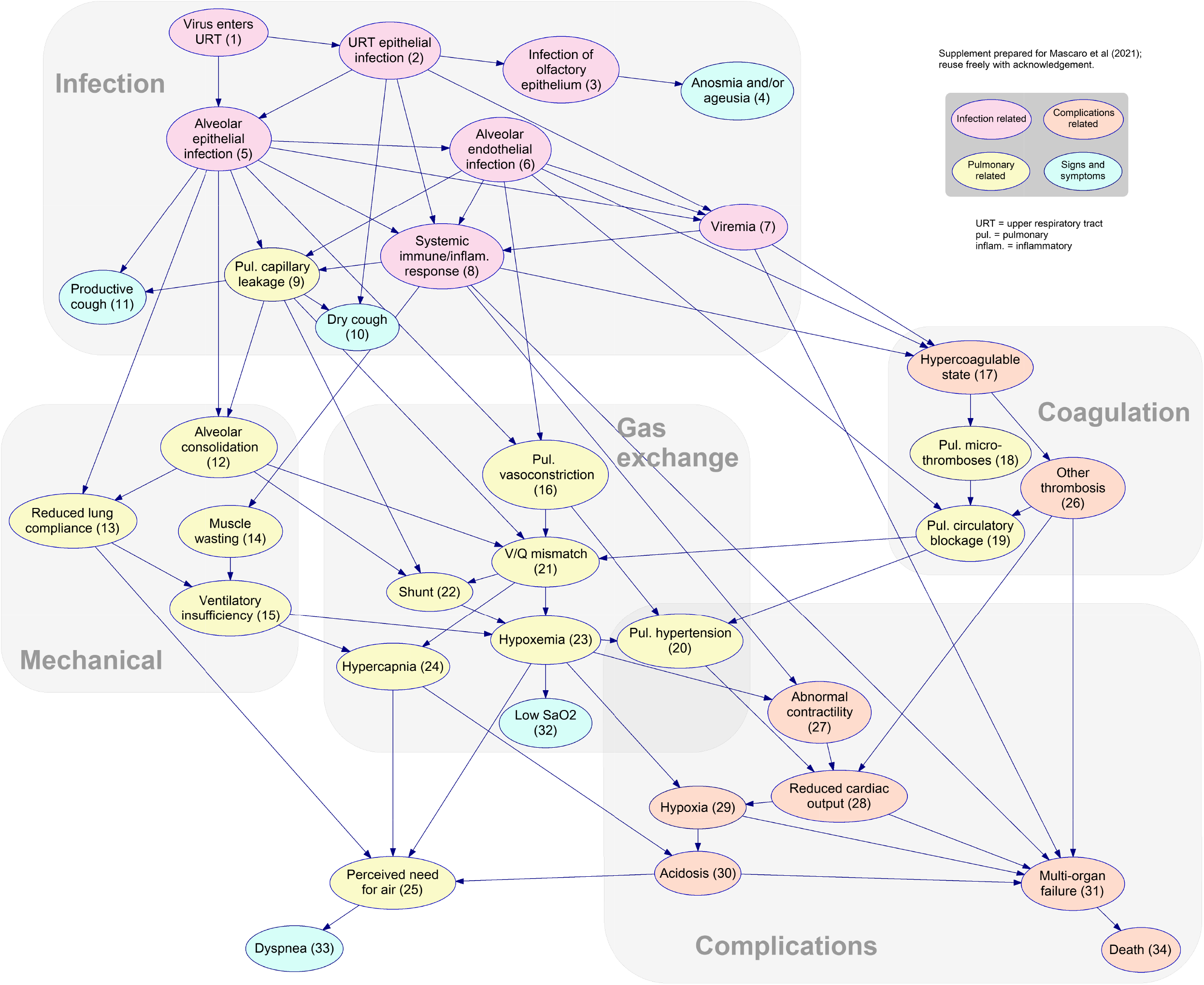
*Respiratory* causal DAG v3.8. This depicts the initial pathophysiological process of SARS-CoV-2 in the respiratory system, outlining multiple and often concurrent pathways from viral infection to key downstream complications such as multi-organ failure. Some variables are latent (i.e., not directly observable) but their probability distributions can be inferred from observable evidence such as clinical signs, symptoms and laboratory measurements, not all of which are shown in the diagram. Many mechanisms described in the BN can be influenced by background factors such as age, sex, and comorbidities, which are also not shown. BNs are acyclic, so feedback loops that may occur as the disease progresses are not included in the diagram. We divide the nodes into four color-coded categories: **Infection** process (pink), **Pulmonary** details (yellow), resulting **Complications** (orange), and a few illustrative examples of **Signs and symptoms** (cyan). Within the pulmonary system, we distinguish (using background boxes) three pathways from **Infection** to possible **Complications**: involving problems with **Mechanical** operation of the lungs, **Gas exchange**, and **Coagulation**.

**S2 Fig.**
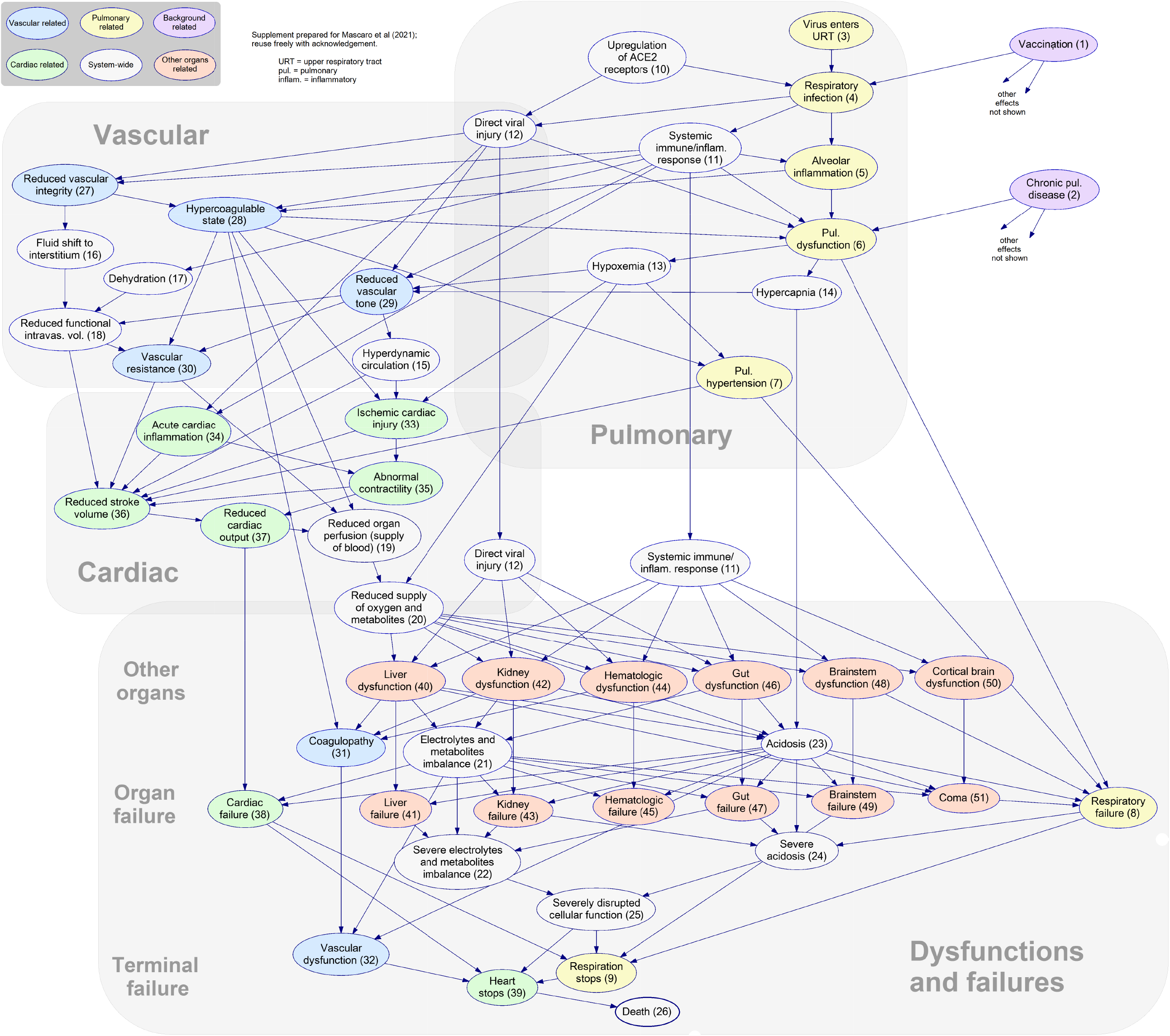
*Complications* causal DAG v3.8. This depicts the main physiological processes underlying the progression of COVID-19 from the initial infection in the **Pulmonary** system (nodes in yellow) to complications in other organs. **Vascular** (nodes in blue) and **Cardiac** (nodes in green) systems are modeled in more detail due to their likely earlier involvement and greater system-wide impact on **Other Organs** (nodes in orange), i.e., liver, kidney, hematologic, gastrointestinal, cortical and brainstem dysfunction. Mechanisms that have a system-wide impact are colored in off-white, and we include two illustrative examples of background factors (nodes in purple).

**S1 Model. *Respiratory* BN v3.8 (indicative parameters only)**. This BN file for the *Respiratory* model may be viewed, manipulated and edited in the GeNIe application (https://www.bayesfusion.com/genie/), which is free for academic use, or reformatted for other BN software. The network structure is depicted in S1 Fig., and in addition the BN includes states for each variable and parameters specifying their influence (which are only rough indications based on our elicitation sessions and literature reviews).

**S2 Model. *Complications* BN v3.8 (indicative parameters only)**. This BN file for the *Complications* model may be viewed, manipulated and edited in the GeNIe application (https://www.bayesfusion.com/genie/), which is free for academic use, or reformatted for other BN software. The network structure is depicted in S2 Fig., and in addition the BN includes states for each variable and parameters specifying their influence (which are only rough indications based on our elicitation sessions and literature reviews).

**S1 Table. *Respiratory* BN dictionary v3.8**. This table for the *Respiratory* BN specifies, for each variable, a description of the variable and its relationships to its parent nodes, supported by references to academic literature listed in a bibliography. Relevant evidence, background factors, and some feedback loops are noted even if not included in the BN diagram.

**S2 Table. *Complications* BN dictionary v3.8**. This table for the *Complications* BN specifies, for each variable, a description of the variable and its relationships to its parent nodes, supported by references to academic literature listed in a bibliography. Relevant evidence, background factors, and some feedback loops are noted even if not included in the BN diagram.

**S3 Table. Members of COVID BN Advisory Group v1.2**. This table lists all the members of our COVID BN Advisory Group who opted to be individually acknowledged, with their institutions and relevant qualifications.

## Acknowledgments

This publication is supported by Digital Health CRC Limited funded under the Australian Commonwealth Government’s Cooperative Research Centres Programme and The Snow Medical Research Foundation. We thank all experts and our COVID BN Advisory Group who participated in the elicitation sessions including surveys, group workshops, and one-on-one meetings. The members of the advisory group are listed in S3 Table. We also thank Dr Sally Shrapnel for helpful feedback on the draft of this article.

---

**S1 Table.**
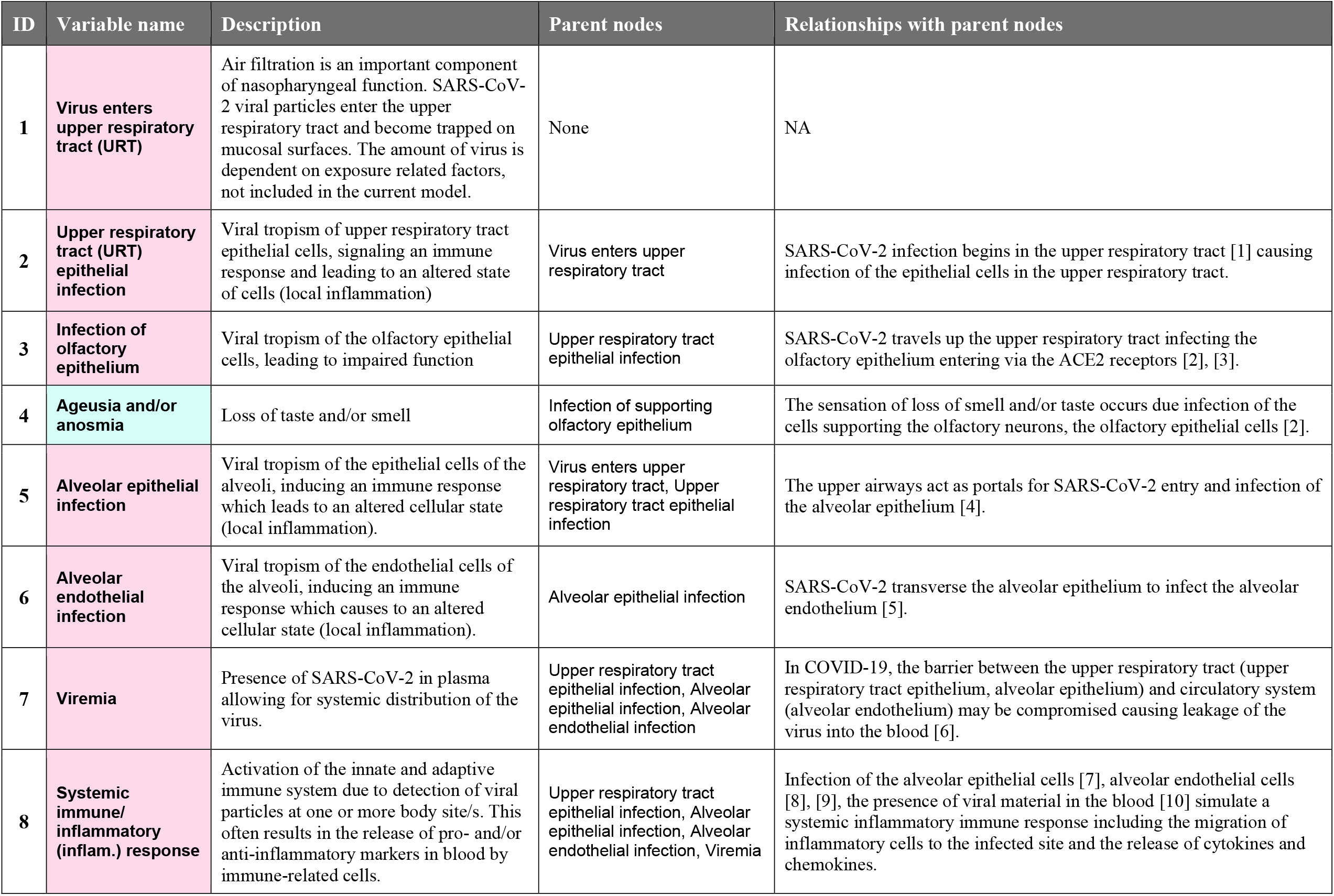
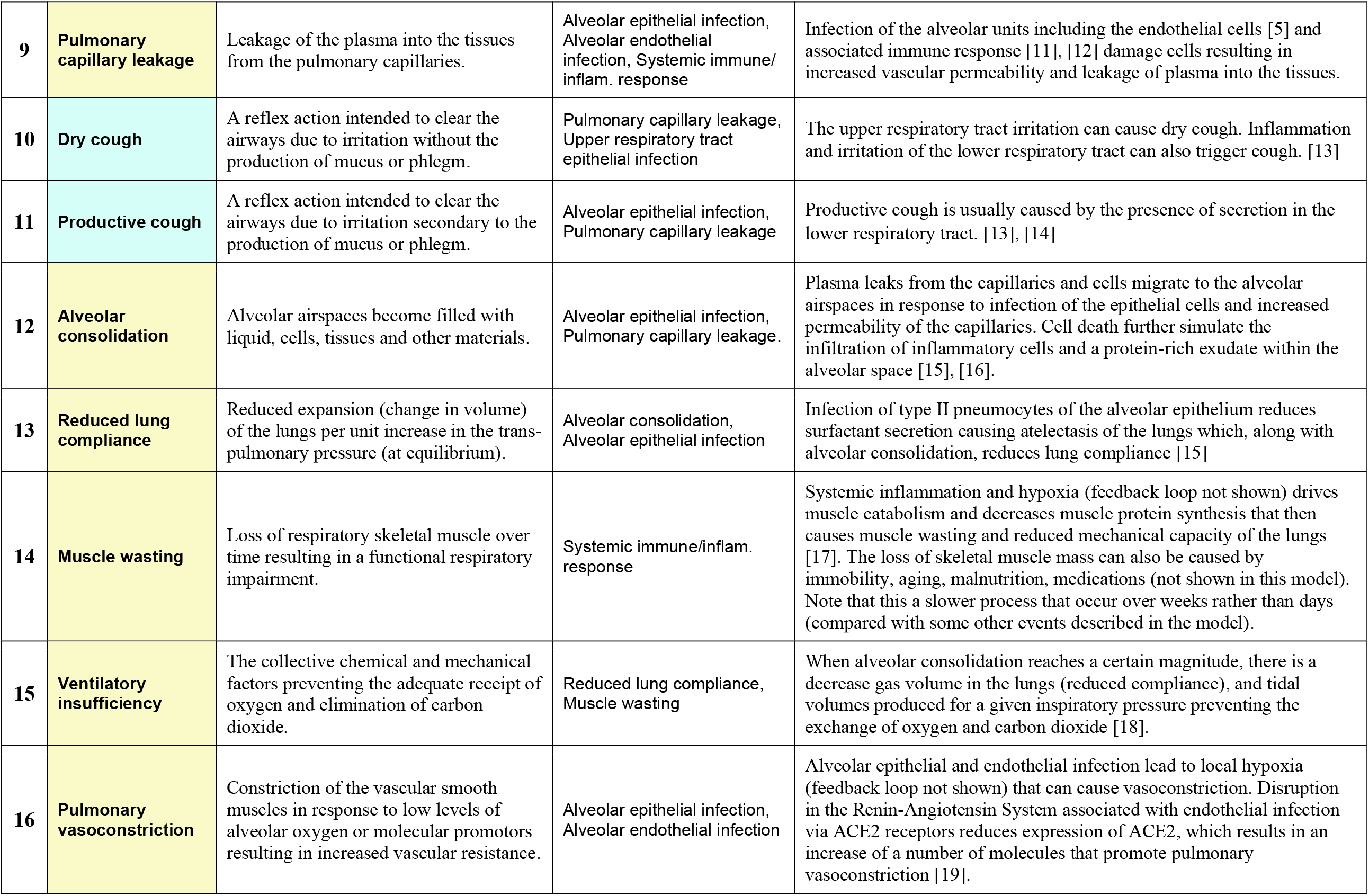
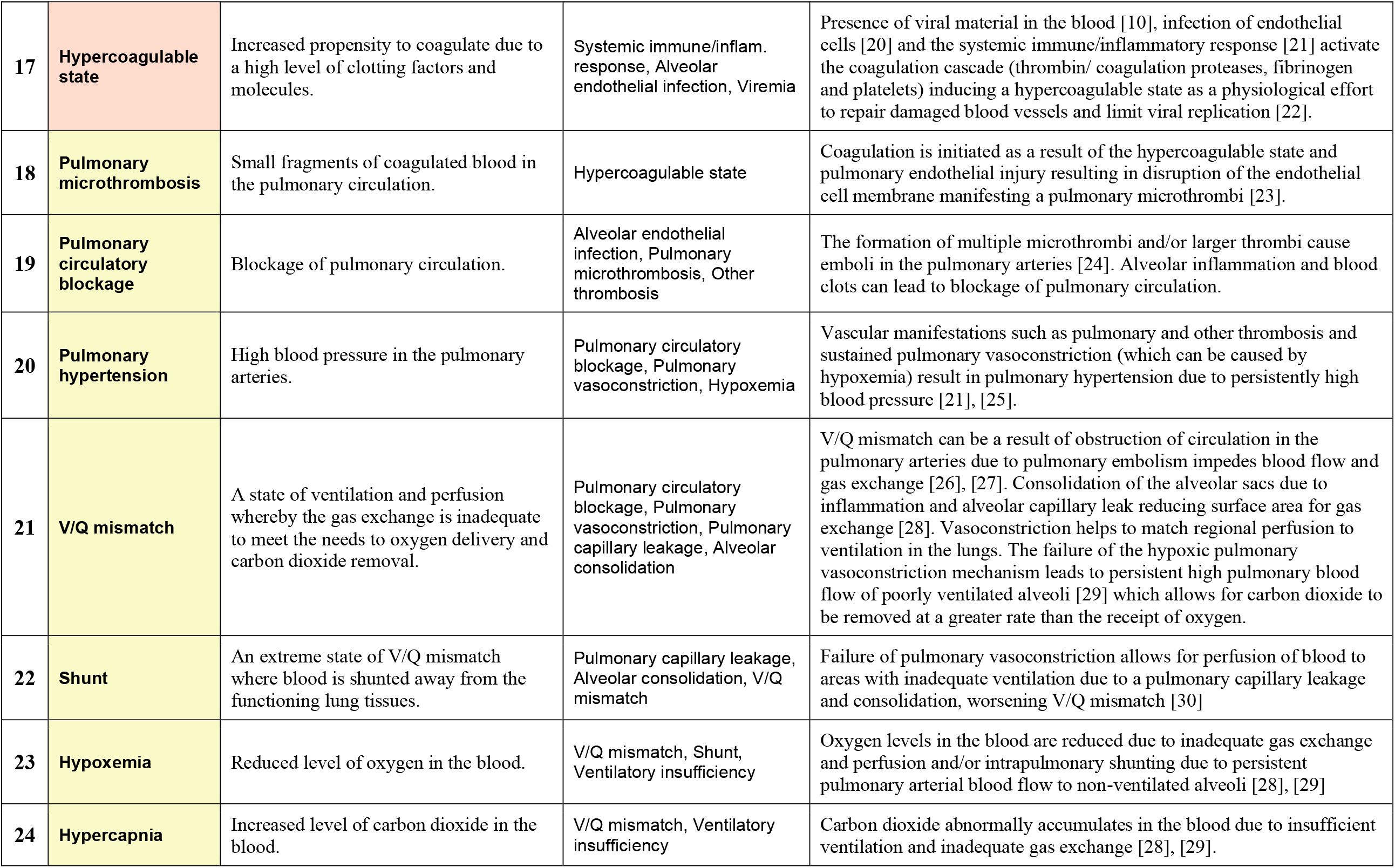
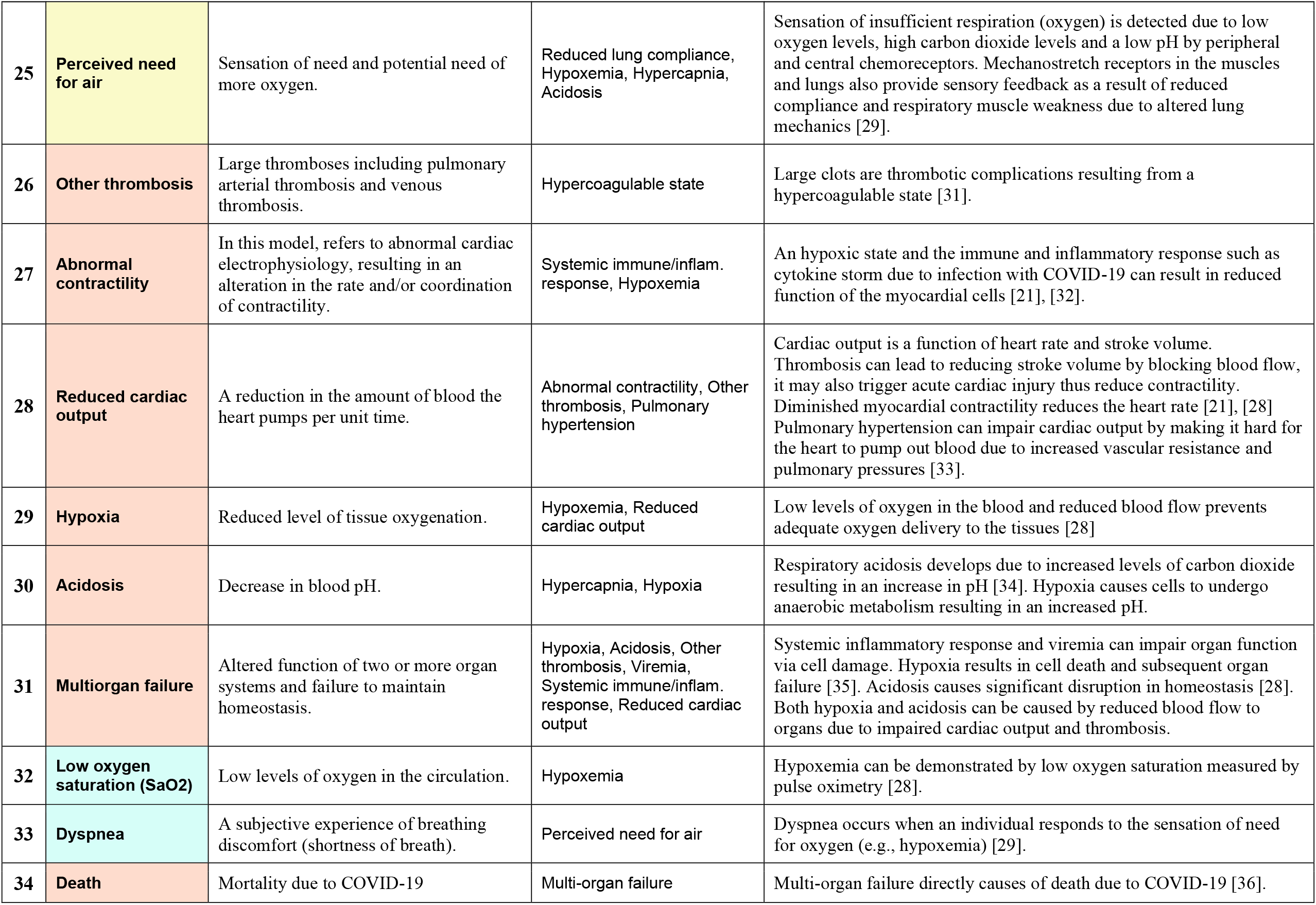
*Respiratory* BN dictionary v3.8. Supplement prepared for Mascaro et al (2021); reuse freely with acknowledgement.

**S2 Table.**
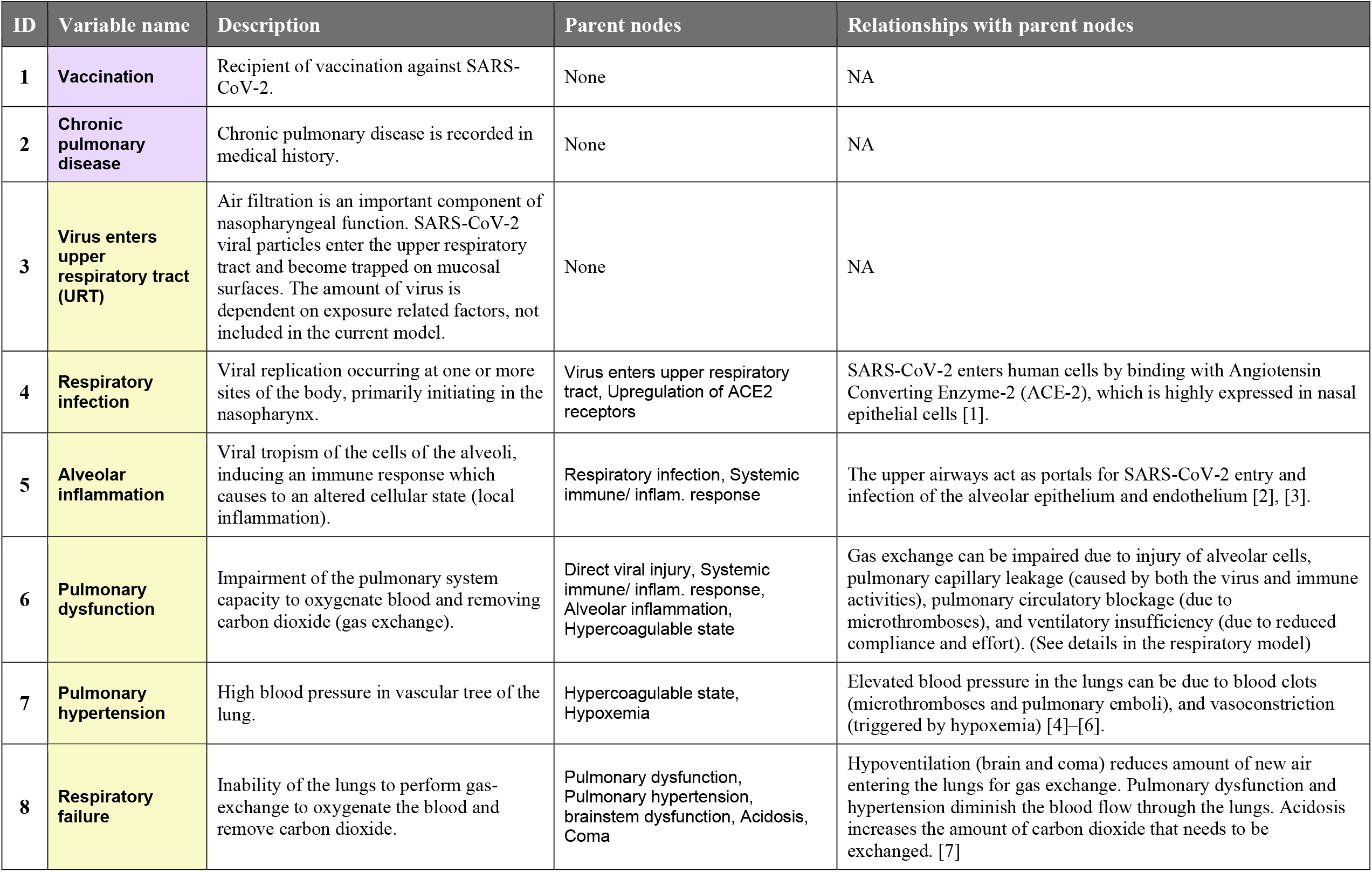
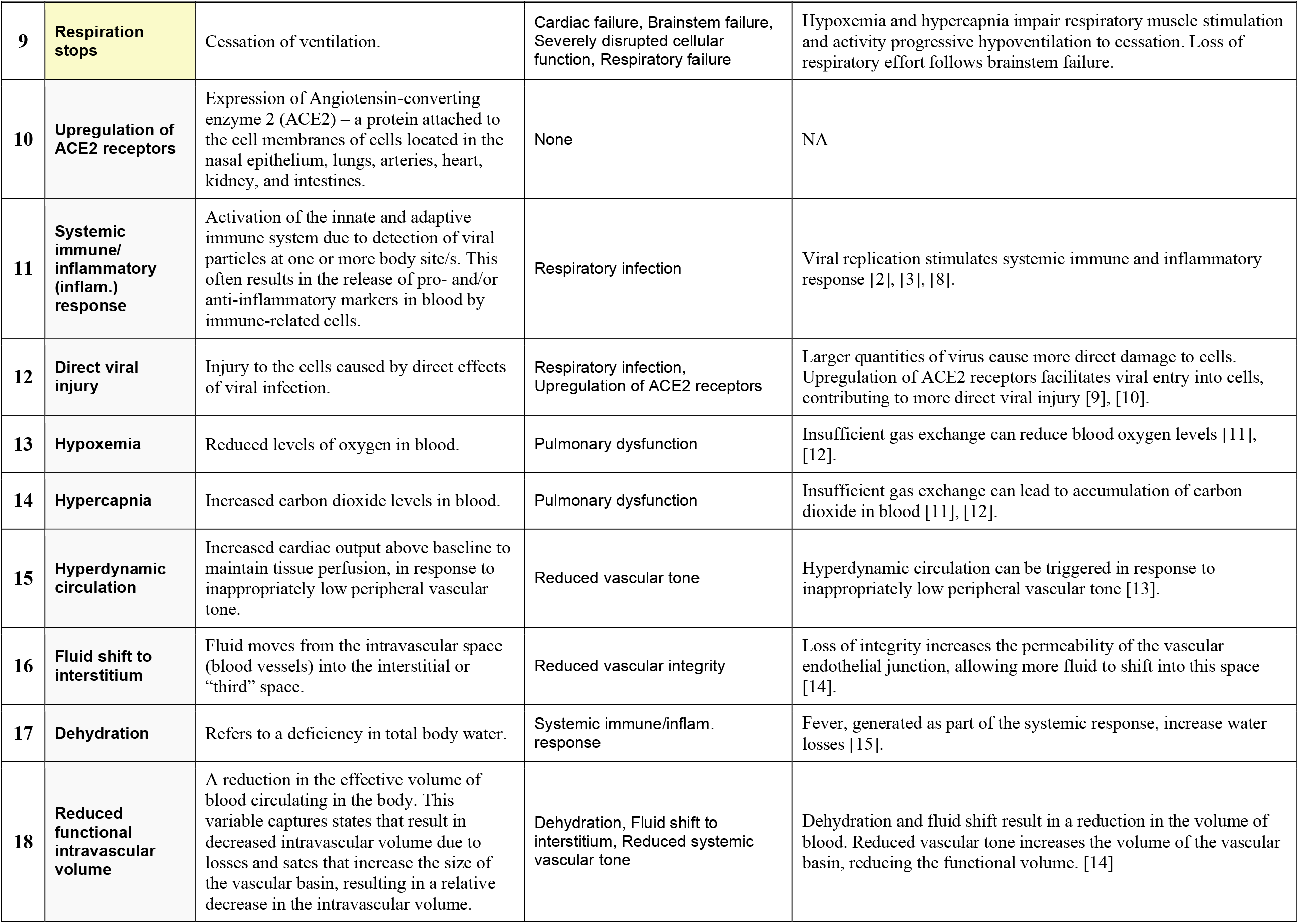
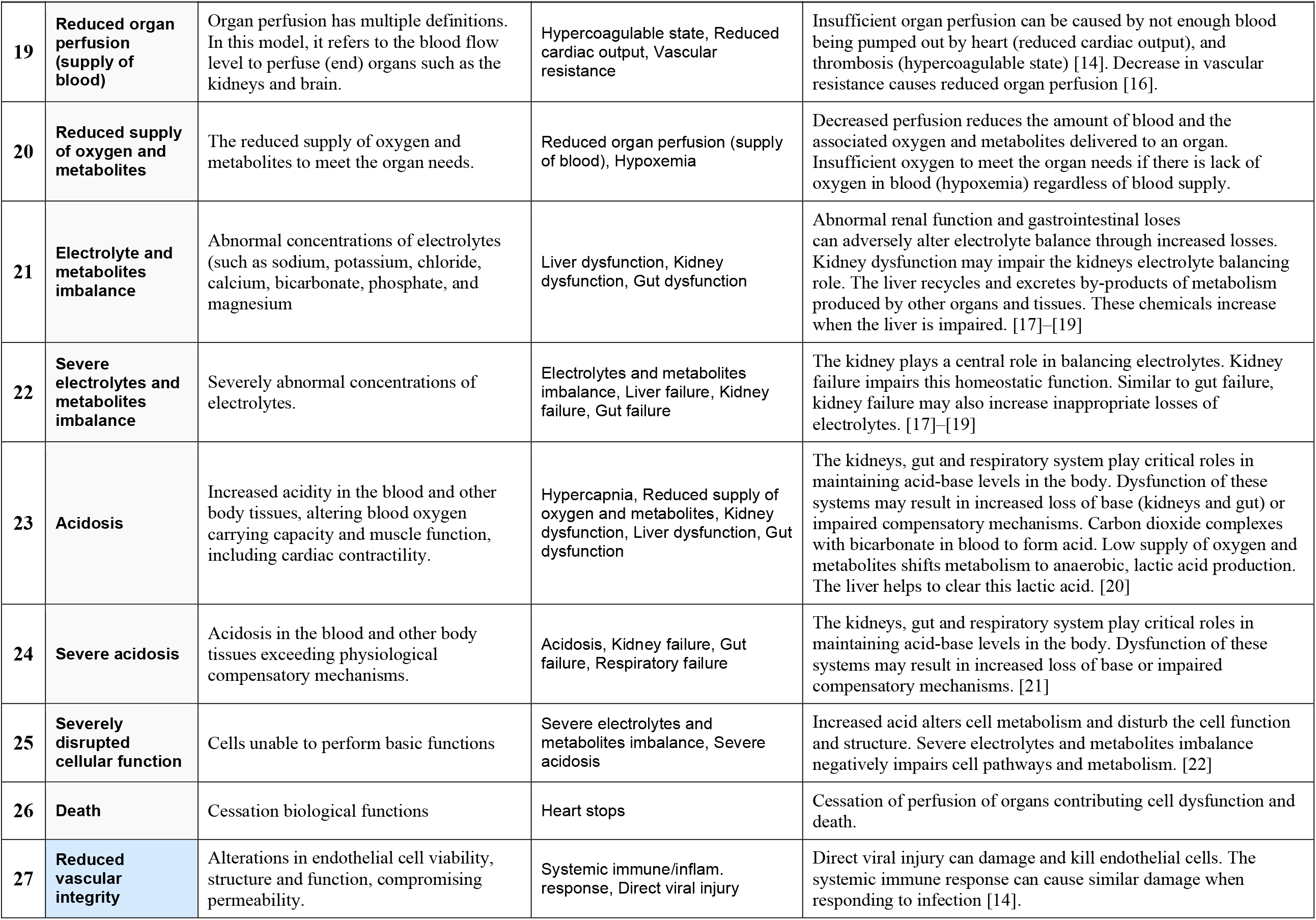
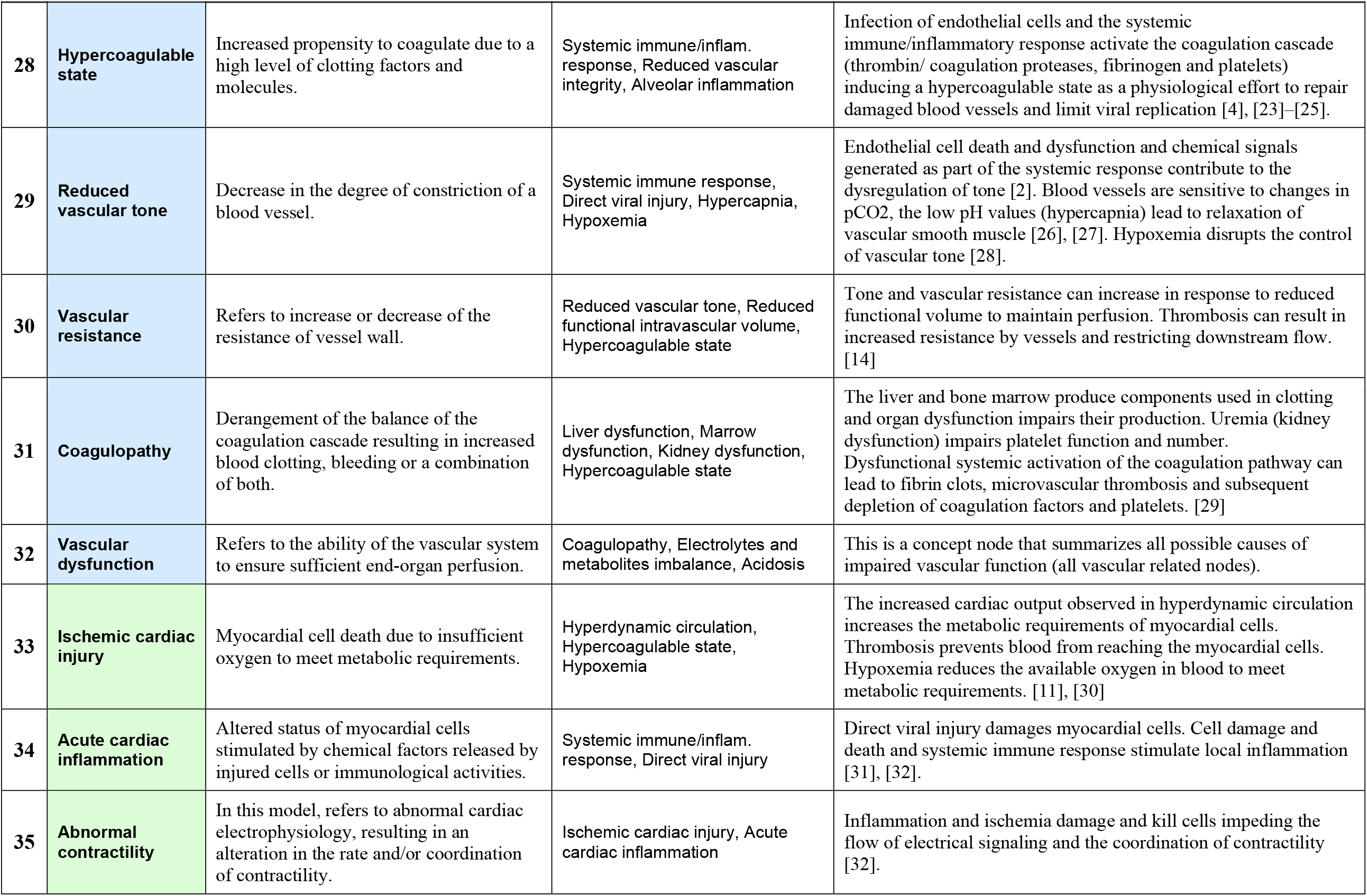
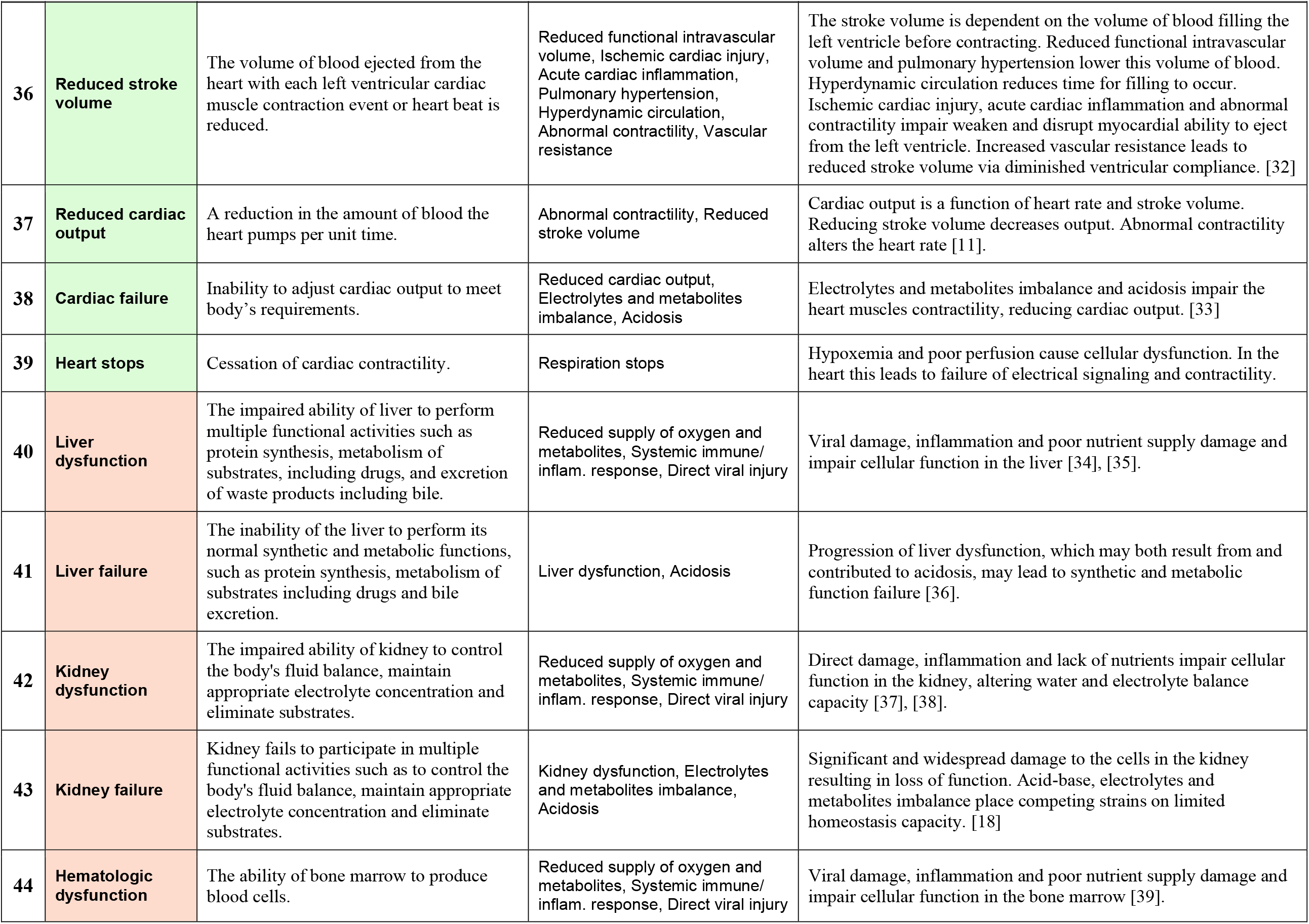
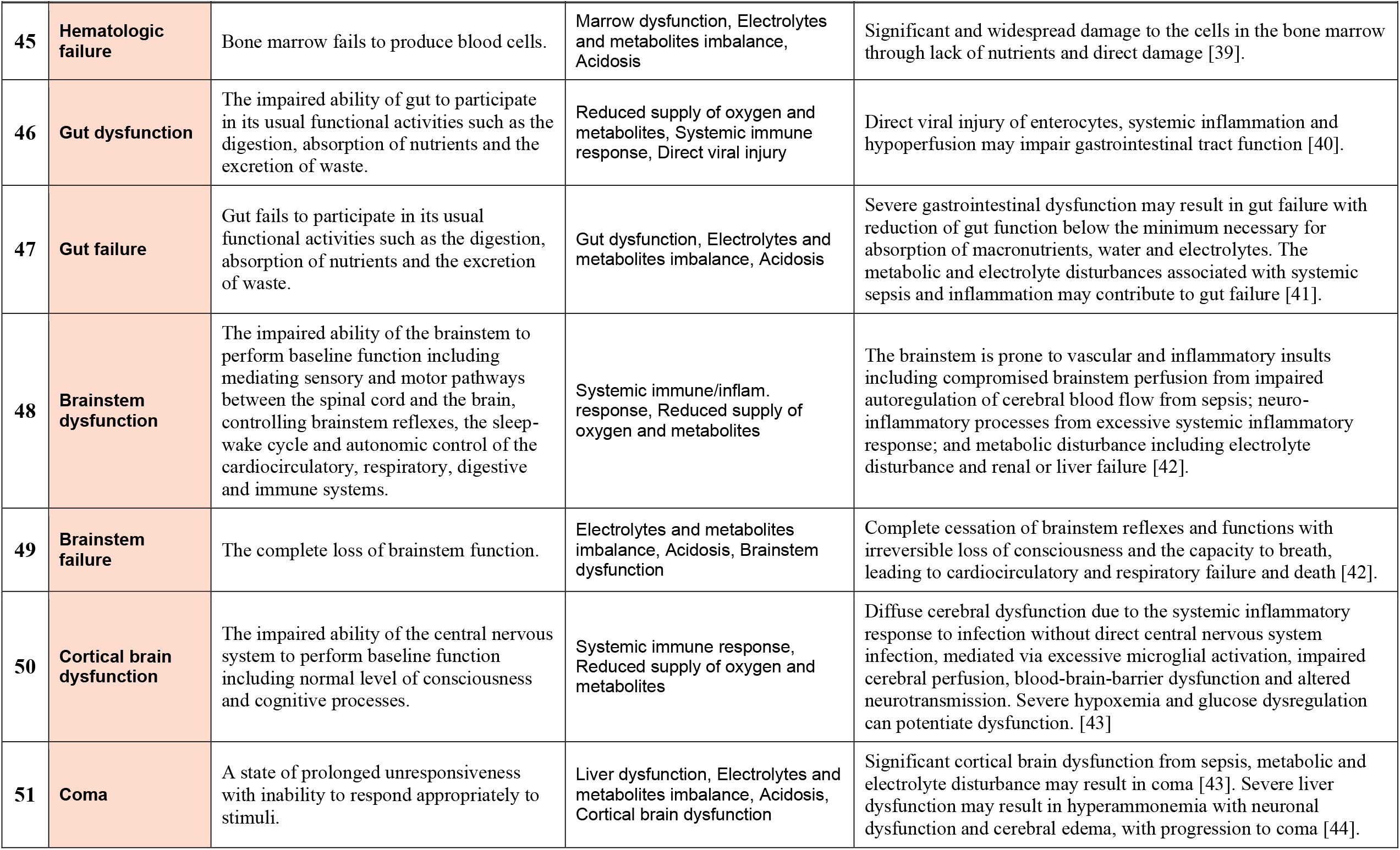
*Complications* BN dictionary v3.8. Supplement prepared for Mascaro et al (2021); reuse freely with acknowledgement.

**S3 Table.**
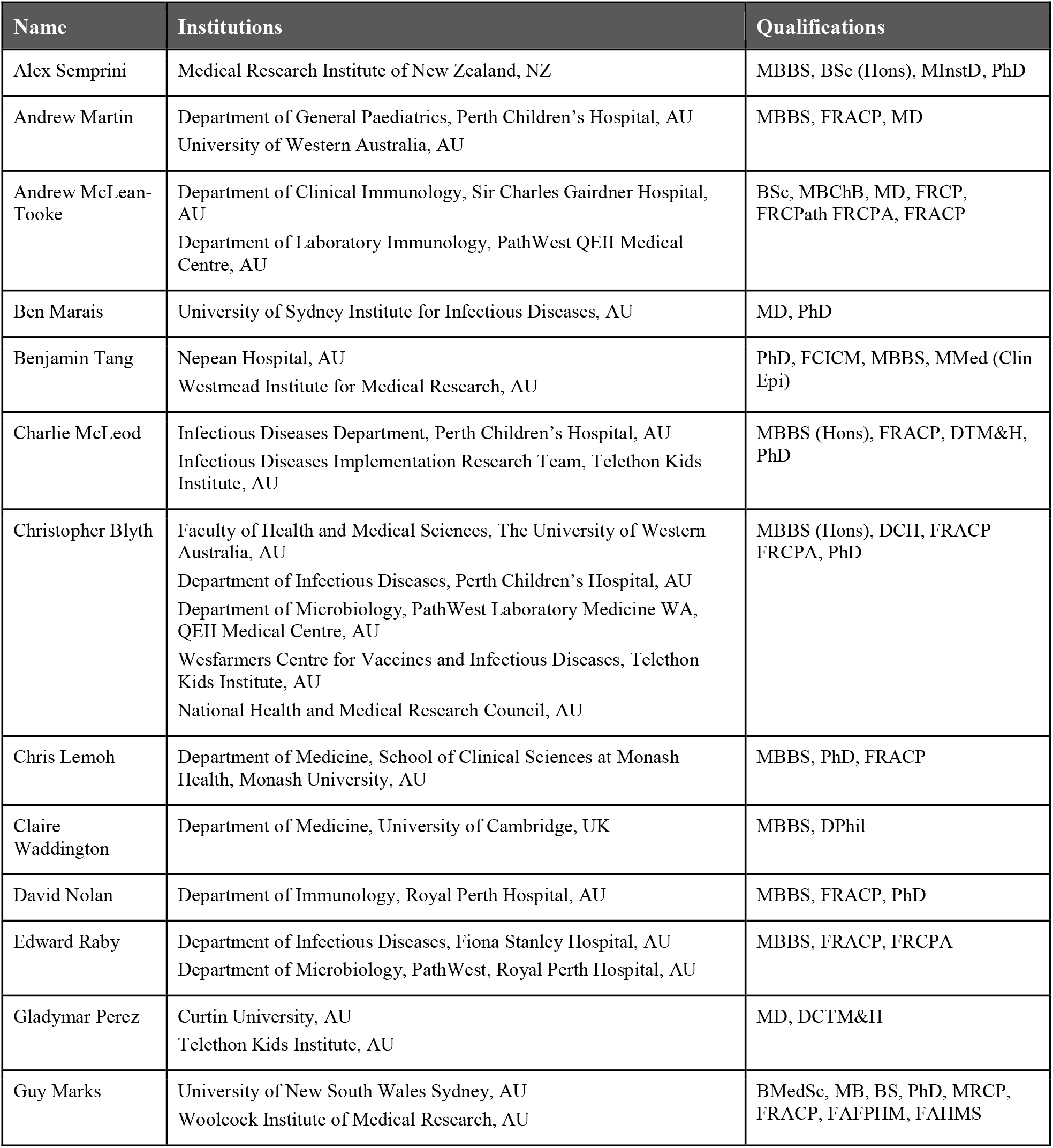
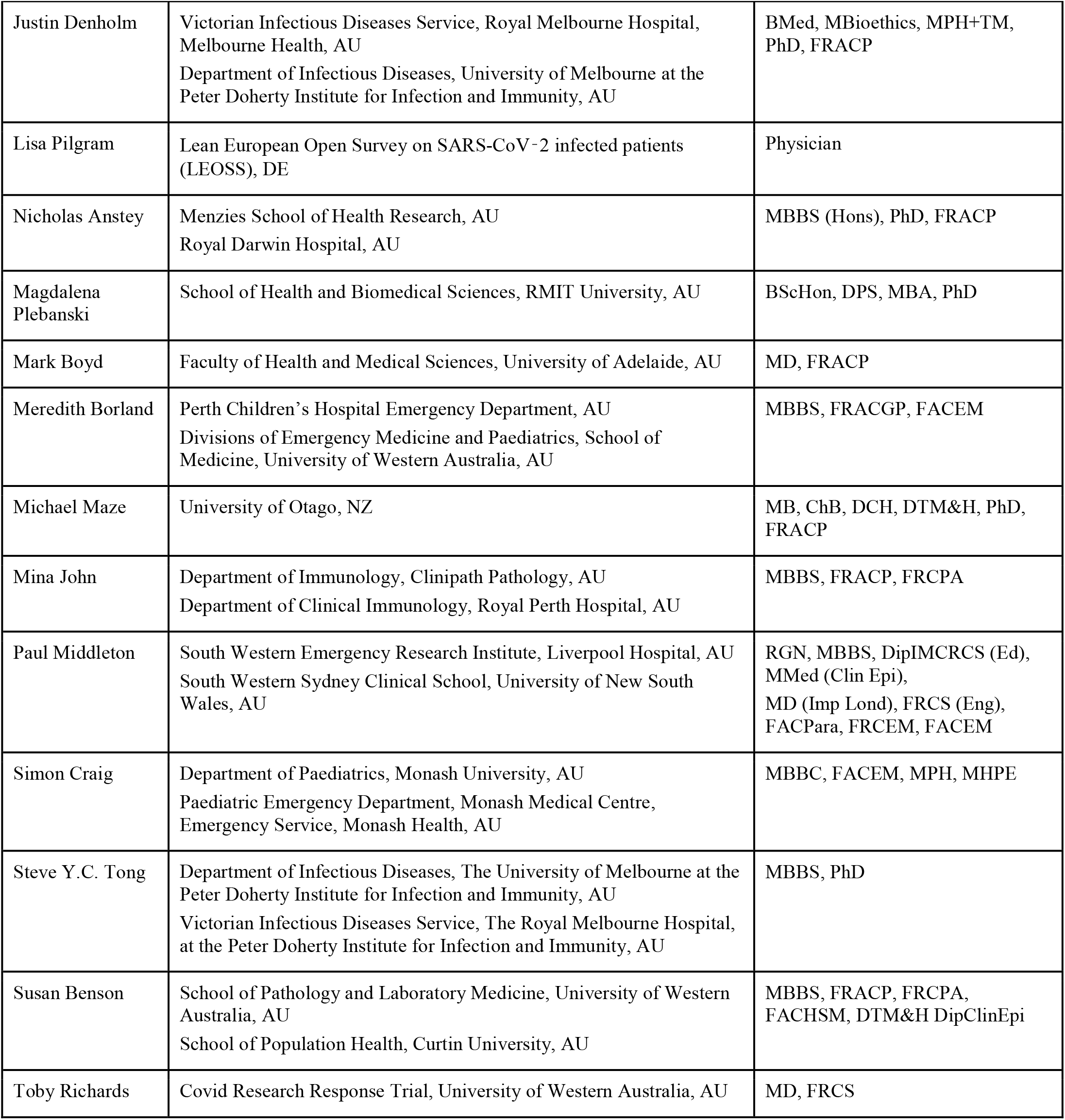
Members of COVID BN Advisory Group v1.2. The following members agreed to be acknowledged individually; others preferred to remain anonymous. We thank all members for participating in the elicitation sessions, including group workshops, one-on-one meetings, and surveys. This supplement was prepared for Mascaro et al (2021); reuse freely with acknowledgement.

## Footnotes

1 In mathematical graph theory, where early work focused on polyhedra, these are traditionally called ‘vertices’ connected by ‘directed edges’; but common and equivalent terms are ‘nodes’ connected by ‘directed links’, ‘arrows’ or ‘arcs’.

2 We used the GeNIe BN software tool (https://www.bayesfusion.com/) to elicit the BNs presented here, and Netica (https://www.norsys.com/) to develop and parameterize subsequent models from datasets. Other widely-used commercial BN software tools include Hugin (https://www.hugin.com/), AgenaRisk (https://www.agenarisk.com/), and BayesiaLab (http://www.bayesia.com/). In addition, research software and tools include Elvira (http://leo.ugr.es/elvira/), R BN libraries (http://www.bnlearn.com/), BNT (https://github.com/bayesnet/bnt/), SamIam (http://reasoning.cs.ucla.edu/samiam/), and BayesPy (https://pypi.org/project/bayespy/).

3 We implemented this procedure in the subsequent development of our parameterized *Progression* DBN. We conducted a survey and group elicitation session dedicated to background factors, where we asked experts which of the *Progression* variables (which do not include all the theoretical variables here) would be affected.

